# Cognitive training in Parkinson’s disease induces local, not global, changes in white matter microstructure

**DOI:** 10.1101/2021.04.23.21255914

**Authors:** Chris Vriend, Tim D. van Balkom, Henk W. Berendse, Ysbrand D. van der Werf, Odile A. van den Heuvel

## Abstract

Previous studies have shown that cognitive training can improve cognitive performance in various neurodegenerative diseases but relatively little is known about the effects of cognitive training on the brain. Here we investigated the effects of our cognitive training paradigm, COGTIPS, on regional white matter microstructure and topology of the structural network. We previously showed that COGTIPS has small, positive effects on processing speed. A subsample of 79 PD patients (N=40 cognitive training group, N=39 active control group) underwent multi-shell diffusion weighted imaging pre- and post-intervention. Our pre-registered analysis plan (osf.io/cht6g) entailed calculating white matter microstructural integrity in five tracts of interest, including the anterior thalamic radiation (ATR) and calculating the topology of the structural connectome. Training-induced changes were analyzed with linear mixed-models. Relative to the active control condition, cognitive training had no effect on network topology. Cognitive training did lead to a reduction in fractional anisotropy in the ATR (B[SE]: -0.32 [0.12], p=0.01). This reduction was associated with faster responses on the ToL task (r = 0.42, P = 0.007), but this just fell short of our statistical threshold (P<0.006). Post-hoc analyses showed that this was not due to changes in fiber density and cross-section, suggesting that that the observed effect in the ATR are due to training-induced alterations in neighboring fibers running through the same voxels, such as intra-striatal and thalamo-striatal fibers. These results indicate that eight weeks of cognitive training do not alter network topology, but can have subtle local effects on structural connectivity.

## INTRODUCTION

Cognitive training (CT) has not only been shown to have a positive effect on cognitive performance in both healthy and diseased populations, it also influences the brain on a circuit and system level. The majority of studies has focused on the effects of CT on functional activation or connectivity, showing that CT can increase neural efficiency and counteract aging- or disease-related neural dysfunction (van Balkom et al. 2020). CT may also seems to have an effect on the microstructure of the white matter and structural connectivity. In healthy elderly, CT increased the integrity (measured by fractional anisotropy (FA)) of the uncinate fascicle [1] and anterior thalamic radiation (ATR) [2]. The effects of CT may even be evident after 12 months, with one study showing maintenance of white matter integrity in the posterior parietal cortex in the CT but not control condition [3]. This study also showed a positive association with training-induced improvement in processing speed. Other studies have found no effect of CT on white matter integrity [4,5], but studies have overall been small. So far only one study has investigated the effects of CT on white matter integrity in Parkinson’s disease (PD), showing no changes immediately after CT [6], or at one year follow-up [7].

Compared with healthy controls, PD patients exhibit lower white matter integrity in the corpus callosum and cingulate and temporal regions [8]. The inferior longitudinal fascicle also seems to be particularly vulnerable to the PD pathology and is associated with cognitive deterioration [9]. Another study also showed that relative to healthy participants, white matter integrity progressively deteriorates from relatively intact in cognitive preserved PD patients, to widespread dysfunction in PD patients with mild cognitive impairment (MCI) and dementia, predominantly in the ATR, corpus callosum, inferior and superior longitudinal fascicles and the cingulum [10]. Using graph analysis, studies have shown that the topology of the structural connectome of PD patients is less efficient and clustered compared with healthy controls [11-14], especially in PD patients with cognitive impairment [15]. No study has yet investigated the effect of cognitive training on the topology of the structural connectome in PD patients.

In the COGnitive Training In Parkinson Study (COGTIPS), we investigated the efficacy of a home-based online CT relative to an active control condition [16]. We included 140 PD patients and acquired MRI scans in a subset of 85 PD patients to assess the effects of CT on the function and structure of the brain. Here we report on the effects of CT on white matter microstructure and the structural connectome. Based on previous studies we expected CT to improve the white matter microstructure of the corpus callosum, inferior longitudinal fascicle and ATR. We additionally hypothesized that CT would improve the topology of the structural connectome, defined as an increase in the global efficiency and average clustering coefficient.

## METHODS

### Participants and intervention

This study is part of the COGTIPS clinical trial [16]. Participants (N=140) were randomized to an experimental CT or an active control in a 1:1 fashion. In both conditions, participants performed an online, home-based, computerized intervention with a duration of eight weeks, three times a week for approximately 45 minutes per session. The experimental CT condition consisted of 13 training games that were adapted from the ‘Braingymmer’ online CT platform (www.braingymmer.com, a product by Dezzel Media). Difficulty of these games was adaptive (i.e. adjusted to performance of the participant) and focused on executive functions, processing speed, attention and visuospatial functions. The AC consisted of three low-threshold games without difficulty adjustment. We acquired an MRI scan from 85 participants. General trial inclusion criteria were 1) mild to moderately advanced idiopathic PD (Hoehn & Yahr stage < 4), 2) significant subjective cognitive complaints (PD Cognitive Functional Rating Scale score > 3), access to and proficiency in using a computer or tablet with internet. General exclusion criteria were 1) a Montreal Cognitive Assessment score < 22, 2) indications of current drug- or alcohol abuse, 3) moderate to severe depressive symptoms (Beck Depression Inventory score > 18), 4) an impulse control disorder, 5) psychotic symptoms except for benign hallucinations, or 6) a history of traumatic brain injury. Exclusion criteria for participation in the MRI study were 1) presence of metal in the body (e.g., a neurostimulator), 2) pregnancy, 4) difficulty lying still for 60 minutes (e.g. due to shortness of breath), 5) a space occupying lesion or 6) significant vascular abnormalities (Fazekas > 1). This study was approved by the medical ethical committee of VU University medical center and performed in accordance with the declaration of Helsinki. All participants provided written informed consent. The trial was registered at clinicaltrials.gov under NCT02920632.

### Image acquisition

MRI scans were acquired on a GE 3.0T Discovery MR750 (General Electronics, Milwaukee, US) equipped with a 32-channel head coil at the Amsterdam UMC location VUmc. We acquired diffusion weighted images with a multi-shell single-spin echo echo-planar imaging sequence (TR = 7350 ms, TE = 81 ms, 2.5×2.5 mm2 in-plane resolution with 56 slices of 2.5 mm; no gap) with 73 interleaved directions (25 b = 1000 s/mm^2^, 24 b = 2000 s/mm^2^ and 24 b = 3000 s/mm^2^) and 7 non-diffusion weighted volumes (b = 0 s/mm^2^). We additionally acquired a 3D T1-weighted structural magnetization-prepared rapid acquisition gradient-echo (MPRAGE) with scan parameters according to the ADNI-3 protocol [17]: TR = 6.9 ms, TI = 900 ms, TE = 3.0 ms, matrix size 256 × 256, 1 mm3 isotropic voxels. Patients followed the same protocol at both time points.

### Image processing

A more detailed account of the (pre)processing pipeline is provided in the supplementary methods and the scripts are available from: github.com/chrisvriend/DWI_processing_COGTIPS. Diffusion images were denoised using the *dwidenoise* tool in MRtrix3 [18] and subsequently processed using the EDDY tool [19] in FMRIB Software Library (FSL) version 6.0.1 [20]. Quality of the DWI was evaluated using EDDY QC [21] and by calculating the median sum of squared error (SSE) of the b1000 tensor fit. These image quality measures (IQMs) were compared across time and groups using the nparLD package in R (version 4.0.2). DWI volumes were visually inspected for residual motion-related artifacts and deleted if necessary. Scans were excluded in case of >3 volumes per shell with motion artifacts. We used FSL DTI-FIT to fit the tensor to the b=1000 s/mm^2^ data to determine fractional anisotropy (FA), mean diffusivity (MD), axial diffusivity (AD) and radial diffusivity (RD) [22]. We used DTI-TK to register the DWI scans to a common space [23] and subsequently performed tract-based spatial statistics (TBSS) [24] to investigate pre-to-post intervention changes in the white matter microstructure of the: genu of the corpus callosum (gCC), splenium of the corpus callosum (sCC), inferior longitudinal fascicle (ILF) and ATR. The corpus callosum ROIs were derived from JHU-ICBM labels (1 mm), while the ILF and ATR were derived from the JHU-ICBM tracts (25%) atlas. We multiplied each tract with the skeletonized mask (thresholded at mean FA > 0.2) and extracted the median value of the FA, MD, AD and RD in the tract. We performed multi-shell anatomically-constrained (probabilistic) tractography (ACT) with 100 million random white matter seeds to construct a tractogram for each participant in MRtrix3 [18]. SIFT2 was applied to improve the accuracy of the reconstructed fibers and reduce false positive connections [25,26]. The resulting tractogram was converted to a 222×222 structural connectivity matrix with 208 cortical areas derived from the Brainnetome atlas and 14 individually segmented subcortical areas with FreeSurfer.

### Graph measures

We calculated graph measures to determine the topology of the structural brain network. We calculated the global efficiency, modularity and average clustering coefficient from individual connectivity matrices. Global efficiency is the inverse of the average path length and provides a measure for the ability of a network to integrate information [27]. The average clustering coefficient quantifies the segregation of nodes in a network, i.e. the tendency of the network to segregate into locally connected nodes to form a specialized subunit. Modularity measures how many modules a network can be divided into. Modules consist of nodes with stronger connections between them compared with nodes outside their module [27].

### Cognitive measures

Participants performed – among other cognitive tests [16] – a self-paced version of the Tower of London (ToL) task on a laptop computer [28] and a paper-and-pencil version of the Stroop Color-Word Test (SCWT) [29]. These tests were performed on the same day as the MRI scans. The ToL covers various executive functions including planning, inhibition and working memory [30] and consists of 100 pseudo-randomized trials with five difficulty levels (task-load S1 to S5) that are scored on accuracy and reaction time. The SCWT is an attention, processing speed and executive function task [29] and requires the participants to read three cards with 100 items as fast as possible. Here we only considered card I (SCWT-I) where participants have to read words as a proxy for processing speed.

### Data analysis

Multivariate mixed model analyses were performed with the four diffusivity measures in each ROI after training as dependent variables, defined condition (CT or active control) as independent variable and included pre-training diffusivity measures as covariates. Diffusivity measures were Z-transformed and MD and RD values were inverted to ensure that higher values on all four diffusivity measures signified better microstructural integrity. We added age, sex and education level as nuisance covariates in separate adjusted models. We additionally performed exploratory whole-brain voxel-wise analyses on the diffusivity measures within a skeleton of the white matter using permutated (10,000) threshold free cluster enhancement (TFCE) and family-wise error (FWE) correction (P<0.05).

The network topological measures were analyzed with univariate linear mixed-models using network measures after training as outcome, the pre-training value as covariate and condition as independent variable. Age, sex and education level were added as nuisance covariates in separate models. The association between changes in DWI-derived measures (diffusivity or network topology) and training-induced changes reaction time on the ToL task or SCWT Card I were analyzed using repeated measures correlations (rmcorr package in R) [31]. These correlations were corrected for multiple comparisons using a D/AP-Sidak adjustment to take into account the mutual correlation between outcome measures [32]. For the analyses on white matter microstructure, with an alpha of 0.05, 20 outcomes (four diffusivity values x five ROIs) and a mutual correlation coefficient of r = 0.31, the adjusted P-value was set at p=0.006 (determined using quantitativeskills.com/sisa/calculations/bonhlp.htm). For the topological analyses, the adjusted P-value was p= 0.027 (alpha = 0.05, 3 outcomes, r = 0.44). We also explored the effect of the training on the connectivity strength between the default mode network (DMN), frontoparietal network (FPN), ventral attention network (VAN) and dorsal attention network (DAN) and their topology using the Yeo network parcellation [33]. Results on the subnetwork level were corrected for multiple comparisons using the False Discovery Rate (FDR; q=0.05). The analysis plan was preregistered at osf.io/cht6g and performed on the intention-to-treat sample only.

## RESULTS

### Demographic and clinical characteristics

From the original 85 PD patients with DWI data, six were excluded for the analyses of WM microstructure and one additional patient was excluded for the graph analysis (see flowchart in Figure 1). Patients in both conditions were adequately matched on all demographic and clinical measures (see Table 1), except for a higher score on the PD-CFRS in the active control group (U = 600.5, P = 0.02). Supplementary table 1 shows the effects of the intervention on performance on the ToL and SCWT in this subsample of 79 PD patients. Compared with the full sample of PD patients [see 34], we observed similar but not statistically significant effect sizes, likely due to the decrease in power.

**Figure 1.**
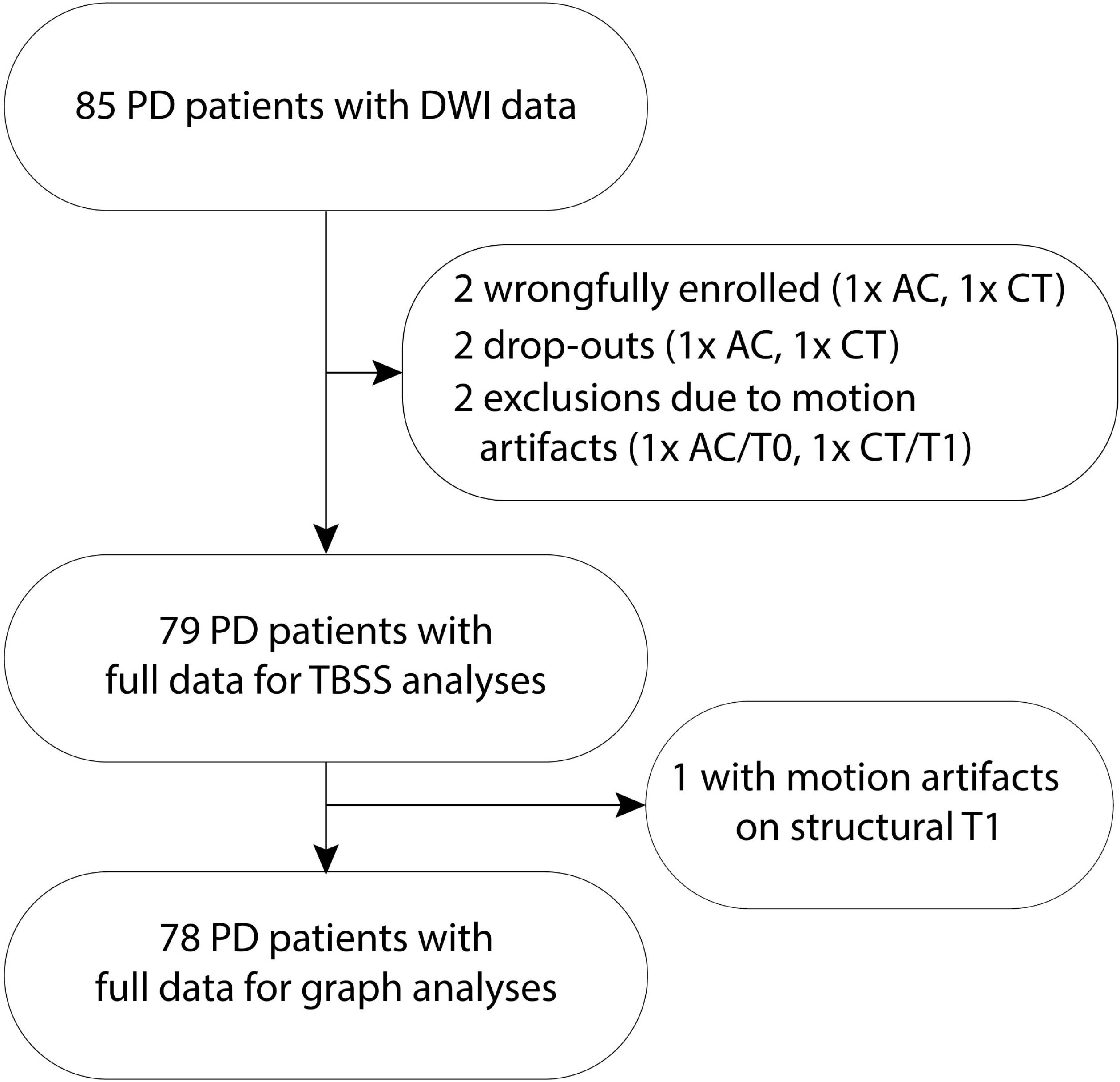
Flowchart. Abbreviations: AC = active control, EXP = experimental cognitive training group, TBSS = tract-based spatial statistics

**Table 1.** demographic and clinical characteristics.

|  | Active control (n=39) | Cognitive training (n=40) | Statistics |
| --- | --- | --- | --- |
| **Sex (N (%))** |  |  |  |
| Male | 26 (66.7%) | 20 (50%) | $\chi^2_{(1)} = 2.3, P = 0.13$ |
| Female | 13 (33.3%) | 20 (50%) |  |
| **Age (years)** | 63.3 (6.4) | 63.3 (8.1) | $t_{(77)} = -0.01, P = 0.99$ |
| **Education (years)** | 17.0 (4.4) | 15.6 (3.6) | $t_{(77)} = 1.5, P = 0.14$ |
| **Education classification (N (%))<sup>†</sup>** | | | $\chi^2_{(4)} = 1.8, P = 0.78$ |
| 3 | - | 1 (2.5%) |  |
| 4 | 3 (7.7%) | 3 (7.5%) |  |
| 5 | 9 (23.1) | 12 (30%) |  |
| 6 | 15 (38.5%) | 12 (30%) |  |
| 7 | 12 (30.8%) | 12 (30%) |  |
| **Disease duration (years, median [range])** | 4 [1-16] | 4 [0-14] | $U = 775, P = 0.96$ |
| **UPDRS-III** | 20.3 (9.4) | 20.6 (8.7) | $t_{(77)} = 0.08, P = 0.94$ |
| **Hoehn & Yahr stage (N (%))** | | | $\chi^2_{(4)} = 3.6, P = 0.47$ |
| 1 | 2 (5.1%) | 3 (7.5%) |  |
| 1.5 | 1 (2.6%) | 5 (12.5%) |  |
| 2 | 19 (48.7%) | 16 (40%) |  |
| 2.5 | 11 (28.2%) | 12 (30%) |  |
| 3 | 6 (15.4%) | 4 (10%) |  |
| **LEDD T0 (median [range] )** | 700 [0-1790] | 682 [80-1665] | $U = 734.5, P = 0.66$ |
| **Medication change (N (%))** | 8 (20.5%) | 7 (17.5%) | $\chi^2_{(1)} = 0.12, P = 0.73$ |
| **LEDD T1 (median [range])** | 734 [0-1790] | 749 [80-1530] | $U = 736, P = 0.66$ |
| **MoCA** | 26.1 (2.4) | 26.5 (1.8) | $t_{(77)} = -0.68, P = 0.50$ |
| **Global cognitive function classification (N (%))** | | | $\chi^2_{(3)} = 4.1, P = 0.25$ |
| Normal cognition | 8 (20.5%) | 12 (30%) |  |
| Single-domain MCI | 5 (12.8%) | 7 (17.5%) |  |
| Multi-domain MCI | 17 (43.6%) | 18 (45%) |  |
| PD dementia | 9 (23.1%) | 3 (7.5%) |  |
| **BDI** | 8.6 (4.3) | 7.9 (4.2) | $t_{(77)} = 0.70, P = 0.49$ |
| **QUIP-RS (median [range]; N=75)** | 20.0 [0-44] | 14.0 [0-60] | $U = 550, P = 0.11$ |
| **PAS** | 11.3 (7.3) | 9.1 (6.2) | $t_{(77)} = 1.4, P = 0.15$ |
| **AS** | 14.0 (4.3) | 13.0 (4.3) | $t_{(77)} = 1.08, P = 0.28$ |
| **Credibility-Expectancy** | 31.4 (6.6) | 33.1 (6.5) | $t_{(77)} = -1.1, P = 0.26$ |
| **PD-CFRS (median [range])** | 10.0 [4-22] | 7 [3.3-18.0] | $U = 514.5, P = 0.009$ |
| **Compliance (% , median [range])** | 100 [70.8-100] | 99.8 [91.9-100] | $U = 779.5, P = 0.99$ |
| **T0-to-T1 interval (days)** | 64.2 (7.2) | 63.8 (4.8) | $t_{(78)} = -.28, P = .78$ |

Data are presented as mean (SD) unless otherwise indicated. <sup>†</sup>According to Verhage education classification.[49] **Abbreviations:** AS = Apathy Scale; BDI = Beck Depression Inventory; PAS = Parkinson Anxiety Scale; PD-CFRS = Parkinson's Disease – Cognitive Functional Rating Scale; LEDD = Levodopa equivalent daily dosage; MCI = mild cognitive impairment; MoCA = Montreal Cognitive Assessment; QUIP-RS = Questionnaire for Impulsive-Compulsive Disorders in Parkinson's Disease – Rating Scale; UPDRS = Unified Parkinson's Disease Rating Scale. For pre-to-post intervention changes in the clinical and cognitive measures see [34]

### Microstructure

Results on the image quality measures are reported in the supplements. There was a significant difference in overall diffusivity in the bilateral ATR between the CT group and the active control group after training while adjusting for the microstructure at baseline (B[SE]: -0.17 [0.08], 95% CI: -0.30 to -0.02, p=0.03; see Table 2 and Figure 2a). This effect was driven by a significant reduction in FA in the CT group after training (B[SE]: -0.32 [0.12], 95% CI: -0.45 to -0.07, p=0.01) that remained significant after additionally adjusting for age, sex and years of education (B[SE]: -0.29 [0.12], 95% CI: -0.53 to -0.05, p=0.02). We also observed a significant difference in MD in the gCC (B[SE]: 0.18 [0.09], 95% CI: 0.006 to 0.35, p=0.04; Figure 2b) but this effect was no longer significant after adjusting for covariates. The other ROIs showed no significant effects (see also supplementary Figure 2). We additionally observed a positive repeated measures correlation between changes in FA in the ATR in the CT group and changes in ToL reaction time (r = 0.42, 95% CI: 0.12 – 0.66, P = 0.007; Figure 3), suggesting that in the CT group responses on the ToL are faster when FA decreases. Nevertheless, this correlation just fell short of our multiple comparisons correction (P_adj_ = 0.006). Whole-brain analyses showed no effects of training (see supplementary results).

**Table 2.**
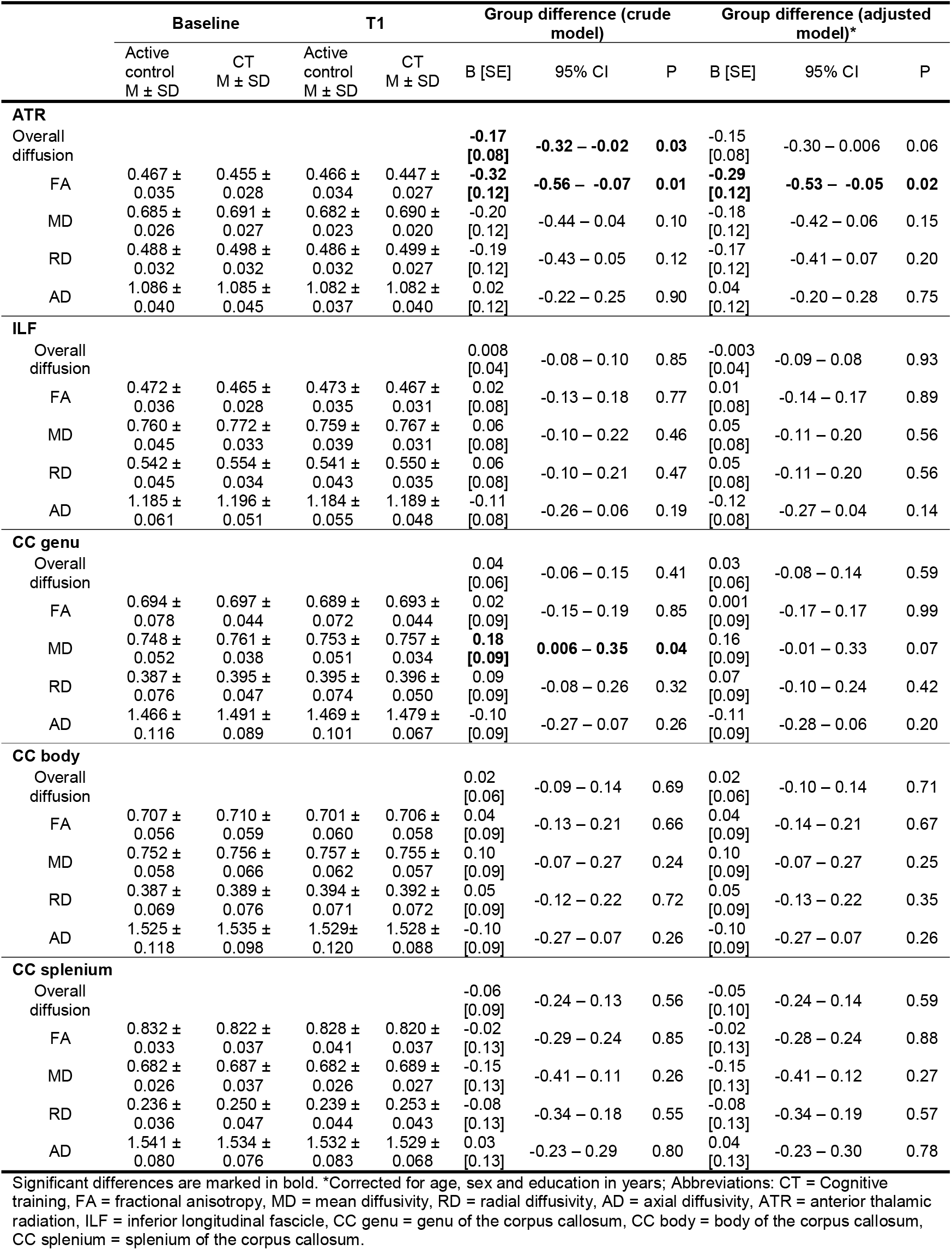
Mixed model analyses of white matter microstructure.

**Figure 2.**
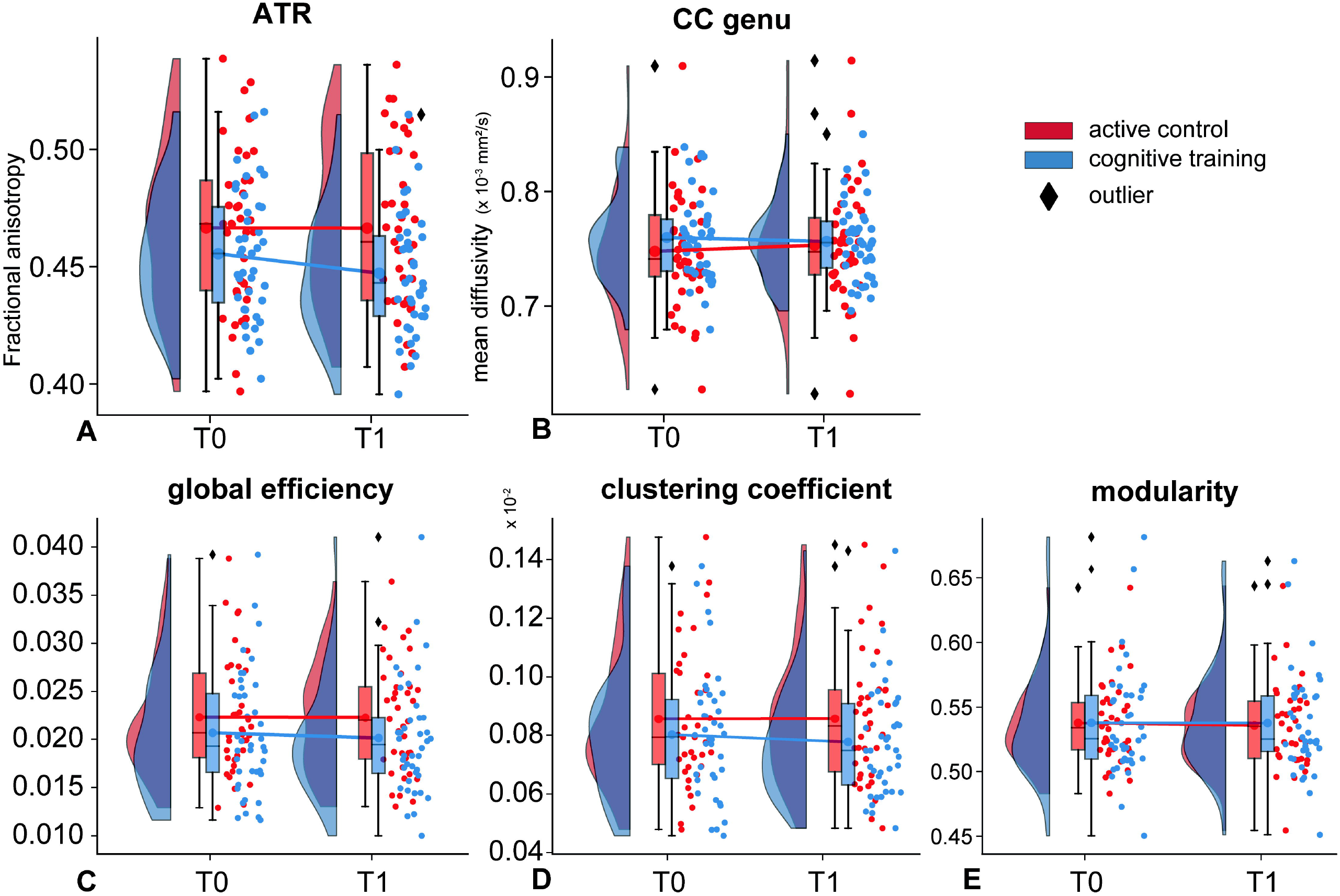
White matter microstructure and network topology. Raincloud plots of the effects of the intervention on diffusivity in the anterior thalamic radiation (a) and genu of the corpus callosum (b) and the network topological measures: global efficiency (c), clustering coefficient (d) and modularity (e). Abbreviations: ATR = anterior thalamic radiation, CC genu = genu of the corpus callosum.

**Figure 3.**
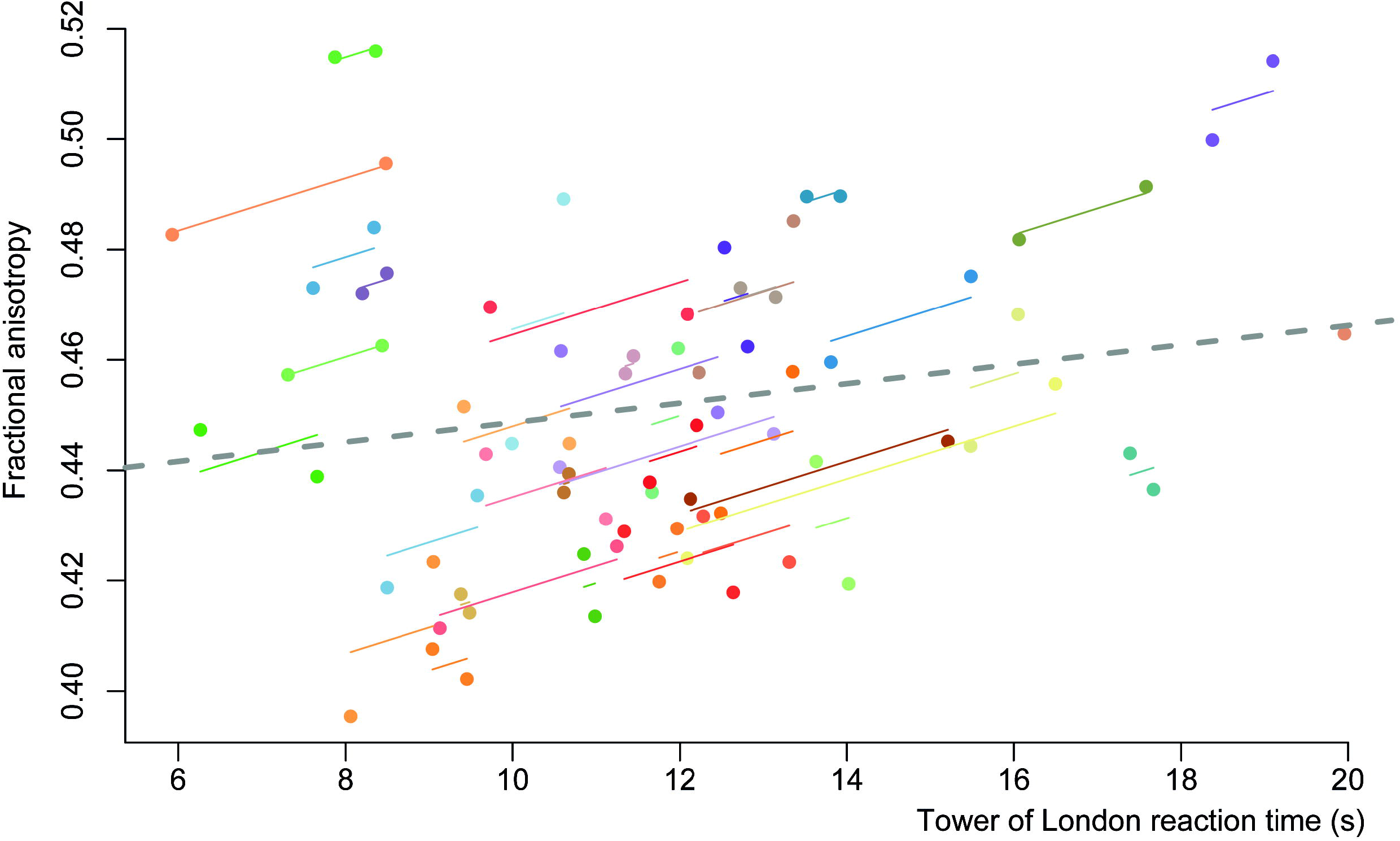
Repeated measure correlations. Correlation plot of the change in reaction time on the Tower of London from pre-training to post-training and change in median fractional anisotropy within the anterior thalamic radiation in the cognitive training group. Note that the FA in the ATR decreased in the CT group, alongside a decrease in (i.e. faster) reaction time.

### Network topology

CT had no effect on whole-brain topology, relative to the active control condition (see Table 3 and Figure 2c-e and supplementary results) and there were no correlations with pre-to-post intervention changes in cognitive performance.

**Table 3.**
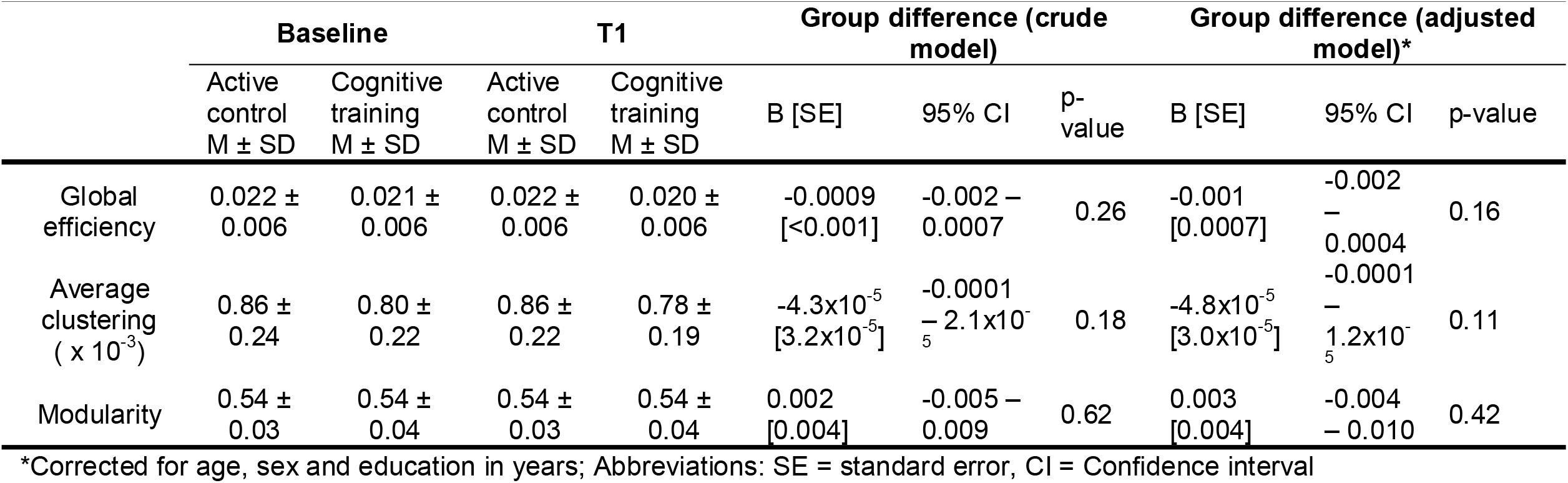
Mixed model analyses of global network topology.

### Post-hoc analyses

We performed post-hoc analyses to follow-up on the observed effects of CT on overall diffusivity – and particularly FA – in the ATR. The effect of training on overall diffusivity was similar across both the left and right ATR, although only the left ATR showed a reduction in FA in the CT group (B[SE]: -0.33 [0.12], 95% CI: -0.56 to -0.09, p=0.008) that remained after adjusting for covariates (B[SE]: -0.30 [0.12], 95% CI: -0.54 to -0.06, p=0.01). We additionally performed a post-hoc ‘fixel’ analysis on the ATR to better understand the unexpected decrease in FA in the CT group. Fixels are *specific* fiber populations within a voxel [35,36]. See supplementary methods for more details. This analyses showed that there were no effects of training on the fiber density or cross-section of the ATR (supplementary Figure 2 and supplementary Table 3). This suggest that the observed reduction in FA in the CT group is not due to changes in density of cross-section of fibers of the ATR.

## DISCUSSION

This study investigated the effects of our online cognitive training program, COGTIPS [16], on WM microstructure and topology of the structural connectome. We showed that in the CT group, relative to an active control condition, overall diffusivity within the ATR decreased, which was driven by a reduction in FA, while MD in the gCC increased. Only the FA reduction in the (left) ATR in the CT group remained significant after correcting for covariates. Interestingly, the decrease in FA in the ATR was associated with faster responses on ToL task (although this fell just short of our multiple comparison threshold). Training-induced faster responses on the ToL task were the main finding on the behavioral level of our randomized controlled trial [34]. CT had no effect on network topology on neither the global or subnetwork level.

The ATR is a major fiber bundle that connects the anterior and midline nuclei of the thalamus with the prefrontal cortex [37]. Due to its trajectory it is critically involved in the associative cortico-striatal-thalamo-cortical (CSTC) circuit and its associated functions [38,39]. Multiple studies have shown dysfunction of the associative CSTC circuit and associated structures in PD, particularly in relation to cognition [40-42]. Based on previous CT studies and the role of the ATR in this circuitry we hypothesized that CT would increase FA relative to the active control group. Surprisingly, our results show an opposite pattern. FA measures the degree of diffusion within a single direction and therefore a decrease in FA may signify either lower axonal density (e.g. due to demyelination and increased free-water diffusion) or a higher proportion of crossing fibers (i.e. diffusion along multiple fibers within a voxel). Standard tensor models cannot disentangle these two possibilities [43] due to averaging the diffusivity of multiple (crossing) fiber populations inside a voxel. We therefore performed a post-hoc analysis of ‘fixels’, i.e. specific fiber populations *within* a voxel [35]. Fixel-based analysis is a relatively new method that allows the quantification of intra-axonal volume by virtue of the density and cross-section of specific tracts, even in voxels that contain crossing fibers [36]. This analysis showed that CT did not alter the intra-axonal volume of the ATR. The ATR runs through the anterior limb of the internal capsule that also contains other fibers. Most of these fibers run in parallel to the ATR (e.g. the superolateral medial forebrain bundle and frontostriatal fibers) [44], but others run perpendicular (i.e. fibers between the caudate nucleus, putamen, pallidum and thalamus; see also supplementary Figure 4). We therefore speculate that the decrease in FA in the CT group is not due to a higher demyelination of the ATR, but due to a higher incidence of crossing fibers that connect these subcortical structures. Unfortunately the resolution of our scans limits our ability to confirm this and necessitates studies at ultra-high field strength [44].

Our results also showed that the reduction in FA in the ATR in the CT group was associated with faster responses on the ToL task. Although this subsample of PD patients with a DWI scan did not show significant differences in cognitive performance after training on the group level, our analyses in the entire sample of 140 PD patients showed positive effects of CT on ToL reaction time, especially for the more cognitively demanding task load 4 [34]. Combined these results are suggestive of training-induced improvement in processing speed during executive functioning that is accompanied with an increase in intra-striatal or thalamo-striatal fibers on the individual level. Although there is some evidence for a role for thalamo-striatal connections in attention [45], the effects of CT on these connections is currently unknown.

The only other but smaller (N=30) study that has investigated the effects of a three month CT paradigm on white matter microstructure in PD patients showed no changes using a whole-brain TBSS approach [6]. Other studies on the effects of CT on white matter microstructure seem to have exclusively been performed in healthy elderly populations and have produced mixed results. Two studies in 12 [5] and 11 healthy elderly [4] showed no effects of training. Conversely, other studies found, relative to a control condition, CT-induced increases in FA in the left ATR [2], increased FA in the left uncinate fascicle [1], or a preservation of left parietal white matter microstructure 12 months after CT [3]. It must be noted that the sample sizes of these studies have overall been small, the studies used different training paradigms and the findings in healthy elderly may not readily be extrapolated to PD or other brain disorders. Interestingly, a recent study in 60 healthy elderly on the effects of a two year multi-domain training program involving not only CT but also diet, exercise and vascular risk management, also observed widespread *decreases* in FA in the intervention group relative to the control group [46]. This unexpected finding was mainly seen in left-sided parietal, callosal and subcortical fibers, including a segment of the ATR, and partly mirrors our own findings. They interpreted the decrease in FA, however, as an intervention-induced reversal of astrocytic hypertrophy and axonal swelling.

CT had no effect on the structural connectome, either at the global or subnetwork level. In fact, network topology remained remarkably stable over time, with test-retest intra-class correlations (ICC) >0.85 for the three global measures. To the best of our knowledge, only one previous study has investigated the effects of a CT on the structural connectome [47] and none have been performed in PD patients. It is possible that the effects of CT are limited to the regional level and do not generalize to global topological changes, at least not within a timeframe of eight weeks. Indeed, Roman and colleagues showed that CT had no effect on global network topology, but they did observe a significant group x time interaction effect on efficiency and strength of a subnetwork involving the temporal, frontal, parietal and insular cortices as well as subcortical areas, using network-based statistics (NBS) [48]. NBS is a statistical approach that identifies a subnetwork based on between-group differences in edge strengths. It is therefore specific to a particular dataset and the identified subnetwork may not necessarily obey the normal hierarchical structure of the brain’s functional systems, such as the ones we investigated here by relying on the Yeo parcellation [33].

A limitation of our study is the lack of a healthy control group that impeded us from assessing the severity of deviating white matter microstructure or topology in our PD patients before training or to assess the potential normalizing effects of CT. Second, although there were no interaction effects for any of the image quality measures, the higher SSE in the active control group may have affected some of our results as the SSE represents the accuracy of the tensor fit (see supplementary results). Lastly, this subsample of PD patients with imaging data was insufficiently powered to detect differences in cognitive performance [see 16]. We did, however, observe similar effects sizes as we did for the full sample. The direction of the repeated measures correlation between ToL reaction time and FA in the ATR is also consistent with the effects observed at the group level, bolstering our findings. Strengths of this study are the large sample size and low attrition, rigorous quality control and a description of the IQMs as well as the use of state-of-the-art registration (DTI-TK) and tractography (MRtrix3) algorithms and the use of a multi-shell DWI sequence to better deal with crossing fibers.

In conclusion, in this largest study on cognitive training in PD patients to date we showed that our CT program, COGTIPS, induces within a timeframe of eight weeks changes in local white matter microstructure that also correlate with cognitive improvement, but has no effect on the topology of the structural connectome at higher levels of organization. Because our post-hoc ‘fixel’ analyses showed no effect on fiber density or cross-section we speculate that the observed changes are due to changes in neighboring (crossing) fibers. These results suggest that CT has subtle and only local effects on structural plasticity.

## Supporting information

COBIDAS checklist

## Data Availability

The datasets generated, used and analyzed during the COGnitive Training In Parkinson Study are available from the corresponding author upon reasonable request.
Code used to proprocess the imaging data is available from github.com/chrisvriend/DWI_processing_COGTIPS

## ACKNOWLEDGMENT

We hereby like to thank Eline Koedijk, Msc. and Monique G.M.S. Schokker, Msc. for their assistance with the visual inspection of the diffusion imaging data, Dr. M.D. Steenwijk for his help with setting up the tractography pipeline and thank drs. A.C.M. Kramer, A.L. Schrijer BSc., drs. A.M. Ticheler, A. van Weert BSc., drs. B. De Azevedo Pinto Castro Maciel, drs. B.E. Olgers, D.N. van Deursen BSc., D.W. van Wylick BSc., drs. E. Koedijk, drs. E.L. Vester, drs. F. Kooij, I. Ashour BSc., drs. I. Zijlstra, J. Breunese BSc., drs. J.F. Stormmesand, drs. J.S.R. Biesbroeck, drs. J.R.C. Verhaegh, K. Basant BSc., drs. L. Drost, drs. M.J. Wagenmakers, M.W. van der Wijk, drs. M.A. Laansma, M.M.A. Schyns BSc., drs. M. Rombouts, drs. M.G.M.S. Schokker, N.M.C. Samoei, BSc., drs. J.P.A. van Dulm MD, R.G.G. Busby, BSc., drs. S. Kasprzak, and V. Joosten, BSc., for their invaluable work on the COGTIPS data collection. This study was funded by the Dutch Parkinson’s Disease Patient Association (grant no. 2015-R04) and the Brain Foundation of the Netherlands (grant no. HA-2017-00227). Part of the participant recruitment was accomplished through Hersenonderzoek.nl, a Dutch online registry that facilitates participant recruitment for neuroscience studies (www.hersenonderzoek.nl). Hersenonderzoek.nl is funded by ZonMw-Memorabel (project no 73305095003), a project in the context of the Dutch Deltaplan Dementie, Gieskes-Strijbis Foundation, the Alzheimer’s Society in the Netherlands and Brain Foundation Netherlands. Participants were additionally recruited through ParkinsonNEXT, a Dutch online registry that aims to unite patients, researchers and clinicians wanting to contribute to research and innovation in Parkinson’s disease and Parkinsonism. ParkinsonNEXT produces information about ongoing studies and facilitates the recruitment of patients.

