## supplement for "Cognitive training in Parkinson’s disease induces local, not global, changes in white matter microstructure"

SUPPLEMENTARY METHODS

### Image preprocessing

Diffusion images were first denoised to improve the generally low signal-to-noise ratio in DWI using the *dwidenoise* tool in MRtrix3 (Tournier *et al.* 2019) (www.mrtrix.org), that exploits the intrinsic redundancy in DWI to remove noise-only principle components from the data without compromising spatial resolution (Veraart *et al.* 2016). Images were subsequently processed using the EDDY tool (Andersson and Sotiropoulos 2016) from FMRIB Software Library (FSL) version 6.0.1 software suite (Smith *et al.* 2004). We estimated the susceptibility-induced off-resonance field using FSL topup from pairs of images with opposite phase-encode directions and fed this field image into FSL EDDY. EDDY corrects the DWI scans for susceptibility-induced distortions, performs within-volume and volume-to-volume motion correction (Andersson *et al.* 2017), and corrects for eddy-current induced distortions and signal drop-out caused by movement during the diffusion encoding (Andersson *et al.* 2016). Quality of the DWI was evaluated using the Quality Assessment for DWI (EDDY QC) tool (Bastiani *et al.* 2019) and by calculating the median sum of squared error (SSE) of the tensor fit in an eroded white matter mask (segmented from the structural T1) as an additional quality measure for the b1000 shell. These image quality measures (IQMs) were compared across time and groups using the nparLD package in R (version 4.0.2) to perform a non-parametric analysis of these longitudinal data. We additionally inspected the volumes (i.e. directions from each shell) of the preprocessed DWI data for residual motion-related artifacts and deleted these volumes where necessary. We allowed a maximum of three volumes per shell to be deleted, otherwise the subject was excluded from the analysis entirely.

### Microstructure

To evaluate training-induced changes on the microstructure of the white matter, we fitted the tensor to the b=1000 s/mm$^2$ data (25 directions and 7 b= 0 s/mm$^2$ volumes) to determine fractional anisotropy (FA), mean diffusivity (MD), axial diffusivity (AD) and radial diffusivity (RD) (Basser *et al.* 1994) using FSL DTIFIT. FA is the most commonly used diffusivity parameter to measure the anisotropy of water molecules within a voxel. A value of 1 represents a uniform direction of diffusion (i.e. along the axis of fibers) and zero unconstrained diffusion (e.g. in CSF). MD measures the total amount of diffusion within the voxel, while AD and RD measure the degree of water diffusion parallel and perpendicular to a fiber tract, respectively. Rather than the FSL default method, we used DTI-TK to register the DWI scans to a common space (Zhang *et al.* 2006). DTI-TK uses the full tensor information (instead of the FA map in FSL) for inter-subject registration and also provides a framework to assess longitudinal changes (Keihaninejad *et al.* 2013). DTI-TK has been shown to better align WM tracts and has a higher registration accuracy than the FSL default and other registration methods (Wang *et al.* 2011; Bach *et al.* 2014). We subsequently performed tract-based spatial statistics (TBSS) (Smith *et al.* 2006) to investigate pre-to-post intervention changes in the microstructure of the white matter of the following regions of interest (ROIs): genu of the corpus callosum (gCC), splenium of the corpus callosum (sCC), inferior longitudinal fascicle (ILF) and ATR. The corpus callosum ROIs were derived from JHU-ICBM labels (1 mm) incorporated in FSL, while the ILF and ATR were derived from the JHU-ICBM tracts atlas with 25% threshold. We multiplied each ROI with the skeletonized mask (thresholded at mean FA > 0.2) and extracted the median value for each of the four diffusivity measures (FA, MD, AD and RD) for data analyses.

**Tractography**

We performed anatomically-constrained (probabilistic) tractography (ACT) implemented in MRtrix3 to construct the structural connectome (Tournier *et al.* 2019). ACT integrates tissue information derived from a segmented structural T1-weighted image to inform the propagation and termination of the streamlines during tractography (Smith *et al.* 2012). The tissue response function was estimated from the preprocessed and bias field corrected multi-shell DWI data (*dwi2response with msm_5tt algorithm*). We subsequently performed Multi-Shell Multi-Tissue Constrained spherical Deconvolution (MSMT-CSD) (Jeurissen *et al.* 2014) to determine the fiber orientation distribution (FODs) in each voxel. ACT was performed by randomly seeding 100 million fibers within the white matter to construct a tractogram. This tractogram was converted to a structural connectivity matrix where each edge represents the number of streamlines between any two brain areas. We created one subject-specific parcellation of the brain for both timepoints by creating a robust template of the T1-weighted images at either timepoint. The robust template was individually parcellated into 224 brain areas: 210 cortical and 14 subcortical areas, using FreeSurfer 6.0.1 software implemented in the fmriprep v1.4.0 pipeline (Esteban *et al.* 2019). The cortical brain areas were derived from registering the Brainnetome atlas to FreeSurfer space and the 14 subcortical areas were individually segmented by FreeSurfer. Because MRtrix3 was unable to reconstruct tracts from area TI of the parahippocampal gyrus (brainnetome regions 117/118) in a majority of participants, these brain areas were excluded from the connectivity matrix, leaving 222 brain areas. In addition, because not every subject's DWI scan fully covered the cerebellum, this structure was not considered in the current analyses. To improve the accuracy of the reconstructed fibers and reduce false positive connections, we performed *Spherical-deconvolution informed filtering of tractograms* (SIFT; *SIFT2 method* in MRtrix3) on the tractogram prior to the generation of the connectivity matrix. This method has been shown to provide a quantitative and biologically meaningful estimation of the structural connectome by applying a weight factor to individual streamlines (Smith *et al.* 2015) and does not warrant further thresholding of the connectivity matrix (McColgan *et al.* 2018).

**Fixel based analysis**

A detailed description of the pipeline can be found here: mrtrix.readthedocs.io/en/latest/fixel_based_analysis/mt_fibre_density_cross-section.html and scripts are available from github.com/chrisvriend/DWI_processing_COGTIPS. We calculated the within-voxel mean fiber density (FD), fiber cross-section (FC) and their combination (FDC) of the skeletonized ATR by non-linearly registering the same tract used for the diffusivity analyses to 'fixel-space'. As with the other reported results, we applied univariate linear mixed-models using these fiber measures after training as outcome and the pre-training value as covariate (condition as independent variable). Intracranial volume was added as nuisance covariate.

SUPPLEMENTARY RESULTS

### Image quality measures

There were no interaction effects for any of the image quality measures, and no group or time effects for relative motion, SNR or CNR for the b1000 shell (see Figure 2). We did, however, observe a significant group effect for CNR for the b2000 ($F(1,\infty)=7.56$, $P= 0.006$) and b3000 ($F(1,\infty)=8.80$, $P= 0.003$) shell and SSE of the b1000 shell (b1000: $F(1,\infty)=10.4$, $P= 0.001$). These group effects were driven by a higher CNR in the CT group compared with the active control group at T1 and higher SSE in the active control group at both time points.

### Explorative analyses

Whole-brain TBSS analyses showed no differences in the white matter microstructure between the two conditions. Explorative analysis of the topology of neurocognitive subnetworks (i.e. efficiency and clustering) and the connections between them also did not reveal any significant effects of training (see supplementary Table 2).

SUPPLEMENTARY TABLES

**Supplementary table 1 – Mixed model analyses of cognitive functioning**

| | Baseline | | T1 | | Group difference (crude model) | | | Group difference (adjusted model)* | | |
|---|---|---|---|---|---|---|---|---|---|---|
| | Active comparator M (SD) | Cognitive training M (SD) | Active comparator M (SD) | Cognitive training M (SD) | B [SE] | 95% CI | p-value | B [SE] | 95% CI | p-value |
| Overall ToL accuracy (%) | 82.946 (7.249) | 80.700 (8.938) | 85.490 (7.934) | 82.310 (11.750) | -0.224 [0.144] | -0.511 to 0.062 | 0.123 | -0.227 [0.138] | -0.501 to 0.047 | 0.104 |
| Task-load (%): S1 | 96.757 (4.119) | 95.481 (5.602) | 97.308 (4.566) | 95.513 (4.973) | -0.282 [0.219] | -0.712 to 0.149 | 0.199 | -0.285 [0.215] | -0.707 to 0.137 | 0.185 |
| S2 | 91.892 (9.155) | 89.972 (10.067) | 94.359 (6.196) | 90.385 (13.098) | -0.307 [0.219] | -0.737 to 0.124 | 0.162 | -0.309 [0.214] | -0.731 to 0.113 | 0.150 |
| S3 | 88.784 (8.612) | 85.935 (12.527) | 89.744 (9.101) | 86.539 (15.818) | -0.193 [0.219] | -0.624 to 0.238 | 0.379 | -0.197 [0.215] | -0.619 to 0.226 | 0.360 |
| S4 | 76.487 (11.599) | 75.573 (14.524) | 81.154 (14.440) | 76.154 (17.414) | -0.284 [0.219] | -0.714 to 0.146 | 0.195 | -0.284 [0.214] | -0.705 to 0.138 | 0.187 |
| S5 | 60.811 (20.967) | 56.539 (18.749) | 64.872 (22.696) | 62.949 (23.971) | -0.055 [0.219] | -0.485 to 0.376 | 0.803 | -0.058 [0.214] | -0.480 to 0.364 | 0.788 |
| Overall ToL reaction time (s) | 12.832 (2.958) | 12.098 (3.211) | 12.519 (3.210) | 11.353 (2.824) | -0.183 [0.118] | -0.418 to 0.052 | 0.125 | -0.148 [0.119] | -0.386 to 0.089 | 0.217 |
| Task-load (s): S1 | 6.424 (1.732) | 5.840 (1.854) | 6.296 (2.463) | 5.206 (1.497) | -0.294 [0.157] | -0.603 to 0.016 | 0.063 | -0.261 [0.158] | -0.572 to 0.050 | 0.100 |
| S2 | 8.279 (2.611) | 7.955 (2.859) | 7.650 (2.040) | 6.942 (2.313) | -0.168 [0.156] | -0.477 to 0.140 | 0.283 | -0.133 [0.157] | -0.443 to 0.177 | 0.399 |
| S3 | 10.908 (2.839) | 10.531 (3.209) | 10.991 (3.674) | 9.682 (3.396) | -0.232 [0.156] | -0.541 to 0.077 | 0.140 | -0.196 [0.157] | -0.507 to 0.114 | 0.213 |
| S4 | 15.673 (4.011) | 15.195 (4.973) | 15.553 (4.800) | 13.880 (5.213) | -0.286 [0.156] | -0.595 to 0.023 | 0.069 | -0.250 [0.157] | -0.561 to 0.060 | 0.113 |
| S5 | 22.876 (5.507) | 20.971 (4.782) | 22.105 (4.715) | 21.159 (4.172) | 0.076 [0.158] | -0.235 to 0.388 | 0.629 | 0.108 [0.159] | -0.205 to 0.422 | 0.496 |
| SCWT I | 55.740 (15.950) | 53.300 (9.310) | 52.640 (12.600) | 50.550 (7.987) | -0.731 [1.702] | -4.118 to 2.657 | 0.669 | -0.226 [1.687] | -3.584 to 3.131 | 0.894 |
| SCWT II | 67.130 (14.351) | 65.230 (12.080) | 67.050 (17.378) | 64.130 (16.010) | -1.012 [2.242] | -5.474 to 3.450 | 0.653 | -1.185 [2.239] | -5.641 to 3.271 | 0.598 |
| SCWT III | 108.030 (40.936) | 114.000 (53.735) | 101.890 (34.654) | 99.250 (24.420) | -5.432 [4.426] | -14.245 to 3.380 | 0.223 | -5.382 [4.498] | -14.336 to 3.572 | 0.235 |
| SCWT interference [(III-II)/II] | 1.598 (0.345) | 1.752 (0.817) | 1.527 (0.202) | 1.561 (0.248) | -0.003 [0.038] | -0.079 to 0.072 | 0.927 | -0.005 [0.039] | -0.083 to 0.073 | 0.904 |

**Supplementary Table 2 – Mixed model analyses of subnetwork topology**

| | **Baseline** | | **T1** | | **Group difference (crude model)** | | | **Group difference (adjusted model)*** | | |
|---|---|---|---|---|---|---|---|---|---|---|
| | Active control M ± SD | Cognitive training M ± SD | Active control M ± SD | Cognitive training M ± SD | B [SE] | 95% CI | p-value | B [SE] | 95% CI | p-value |
| **SUBNETWORK** | | | | | | | | | | |
| DMN efficiency (x $10^{-2}$) | 1.09 ± 0.40 | 0.98 ± 0.33 | 1.11 ± 0.38 | 0.94 ± 0.31 | -0.0008 [0.0005] | -0.002 – 0.0002 | 0.11<br>0.24 | -0.0009 [0.0005] | -0.002 – 0.0001 | 0.08<br>0.15 |
| FPN efficiency (x $10^{-2}$) | 2.10 ± 0.93 | 1.66 ± 0.65 | 2.02 ± 0.82 | 1.57 ± 0.57 | -0.001 [0.0009] | -0.003 – 0.0005 | 0.18<br>0.24 | -0.001 [0.0008] | -0.03 – 0.0003 | 0.11<br>0.15 |
| DAN efficiency (x $10^{-2}$) | 1.64 ± 0.51 | 1.58 ± 0.69 | 1.64 ± 0.54 | 1.51 ± 0.62 | -0.0008 [0.0008] | -0.002 – 0.0009 | 0.36<br>0.36 | -0.0009 [0.0008] | -0.003 – 0.0007 | 0.27<br>0.27 |
| VAN efficiency (x $10^{-2}$) | 0.95 ± 0.28 | 0.91 ± 0.33 | 0.97 ± 0.30 | 0.88 ± 0.32 | -0.0006 [0.0004] | -0.001 – 0.0002 | 0.12<br>0.24 | -0.0006 [0.0004] | -0.001 – 8.7 x$10^{-5}$ | 0.09<br>0.15 |
| DMN clustering (x $10^{-3}$) | 0.77 ± 0.23 | 0.70 ± 0.20 | 0.77 ± 0.22 | 0.68 ± 0.17 | -5.0 x$10^{-5}$ [2.9 x$10^{-5}$] | -0.0001 – 9.2 x$10^{-6}$ | 0.10<br>0.20 | -5.4 x$10^{-5}$ [2.7 x$10^{-5}$] | -0.0001 – 1.5 x$10^{-7}$ | 0.05<br>0.10 |
| FPN clustering (x $10^{-3}$) | 1.03 ± 0.30 | 0.92 ± 0.28 | 1.03 ± 0.28 | 0.89 ± 0.23 | -6.4 x$10^{-5}$ [3.7 x$10^{-5}$] | -0.0001 – 9.3 x$10^{-6}$ | 0.09<br>0.20 | -7.2 x$10^{-5}$ [3.4 x$10^{-5}$] | -0.0001 - -5.2 x$10^{-6}$ | 0.04<br>0.10 |
| DAN clustering (x $10^{-3}$) | 1.05 ± 0.30 | 0.98 ± 0.30 | 1.04 ± 0.28 | 0.96 ± 0.27 | -3.8 x$10^{-5}$ [4.3 x$10^{-5}$] | -0.0001 – 4.8 x$10^{-5}$ | 0.38<br>0.38 | -4.4 x$10^{-5}$ [4.1 x$10^{-5}$] | -0.0001 – 3.7 x$10^{-5}$ | 0.28<br>0.28 |
| VAN clustering (x $10^{-3}$) | 0.77 ± 0.22 | 0.71 ± 0.19 | 0.77 ± 0.20 | 0.69 ± 0.17 | -3.2 x$10^{-5}$ [2.5 x$10^{-5}$] | -8.2 x$10^{-5}$ – 1.9 x$10^{-5}$ | 0.22<br>0.29 | -3.5 x$10^{-5}$ [2.3 x$10^{-5}$] | -8.2 x$10^{-5}$ – 1.1 x$10^{-5}$ | 0.13<br>0.17 |
| DMN – FPN connectivity (x $10^{-3}$) | 4.2 ± 1.7 | 3.5 ± 1.3 | 4.2 ± 1.6 | 3.4 ± 1.2 | -0.0003 [0.0002] | -0.0008 – 9.6 x$10^{-5}$ | 0.13<br>0.28 | -0.0004 [0.0002] | -0.0008 - -7.9 x$10^{-7}$ | 0.05<br>0.16 |
| DMN – DAN connectivity (x $10^{-3}$) | 2.5 ± 0.9 | 2.2 ± 0.9 | 2.5 ± 0.8 | 2.2 ± 0.8 | -0.0001 [0.0001] | -0.0004 – 0.0002 | 0.42<br>0.50 | -0.0001 [0.0001] | -0.0004 – 0.0001 | 0.32<br>0.36 |
| DMN – VAN connectivity (x $10^{-3}$) | 3.2 ± 1.0 | 2.9 ± 0.8 | 3.3 ± 1.0 | 2.8 ± 0.8 | -0.0002 [0.0001] | -0.0004 – 5.2 x$10^{-5}$ | 0.12<br>0.28 | -0.0002 [0.0001] | -0.0004 – 2.5 x$10^{-5}$ | 0.08<br>0.16 |
| FPN – DAN connectivity (x $10^{-3}$) | 6.9 ± 2.0 | 6.2 ± 2.3 | 6.9 ± 2.2 | 6.2 ± 2.2 | -0.0001 [0.0003] | -0.0008 – 0.0005 | 0.60<br>0.60 | -0.0003 [0.0003] | -0.0008 – 0.0003 | 0.36<br>0.36 |
| FPN – VAN connectivity (x $10^{-3}$) | 3.2 ± 1.2 | 2.8 ± 1.0 | 3.2 ± 1.2 | 2.7 ± 1.0 | -0.0002 [0.0001] | -0.0005 – 7.2 x$10^{-5}$ | 0.14<br>0.28 | -0.0002 [0.0001] | -0.0005 – 2.3 x$10^{-5}$ | 0.07<br>0.16 |
| DAN – VAN connectivity (x $10^{-3}$) | 2.2 ± 0.8 | 2.0 ± 0.7 | 2.1 ± 0.8 | 1.9 ± 0.7 | -8.4 x$10^{-5}$ [9.2 x$10^{-5}$] | -0.0003 – 9.9 x$10^{-5}$ | 0.37<br>0.50 | -9.1 x$10^{-5}$ [9.1 x$10^{-5}$] | -0.0002 – 8.9 x$10^{-5}$ | 0.32<br>0.36 |

*Corrected for age, sex and education in years; Abbreviations: DMN = Default mode network, FPN = frontoparietal network, DAN = dorsal attention network, VAN = ventral attention network. Underlined P-values are FDR corrected.

| Supplementary Table 3 – Mixed model analyses of fiber density and cross-section | | | | | | | | | | |
|---|---|---|---|---|---|---|---|---|---|---|
| | **Baseline** | | **T1** | | **Group difference (crude model)** | | | **Group difference (adjusted model)*** | | |
| | Active control M ± SD | Cognitive training M ± SD | Active control M ± SD | Cognitive training M ± SD | B [SE] | 95% CI | p-value | B [SE] | 95% CI | p-value |
| **ATR** | | | | | | | | | | |
| Fiber density | 0.57 ± 0.05 | 0.57 ± 0.03 | 0.57 ± 0.04 | 0.57 ± 0.03 | -0.0006 [0.005] | -0.009 – 0.008 | 0.90 | -0.0002 [0.005] | -0.010 – 0.009 | 0.96 |
| Fiber Cross-section | -0.02 ± 0.09 | -0.05 ± 0.08 | -0.02 ± 0.10 | -0.05 ± 0.08 | 0.002 [0.004] | -0.005 – 0.009 | 0.63 | 0.0009 [0.005] | -0.008 – 0.011 | 0.84 |
| Fiber Density x Cross-section | 0.56 ± 0.08 | 0.55 ± 0.07 | 0.57 ± 0.08 | 0.55 ± 0.06 | $-7 \times 10^{-5}$ [0.006] | -0.013 – 0.013 | 0.99 | $-7 \times 10^{-5}$ [0.006] | -0.015 – 0.015 | 0.99 |

Models are corrected for intracranial volume (ICV). * additionally corrected for age, sex and education in years; Abbreviations: DMN = Default mode network, FPN = frontoparietal network, DAN = dorsal attention network, VAN = ventral attention network. NB: P-values are not corrected for multiple comparisons.

### SUPPLEMENTARY FIGURES

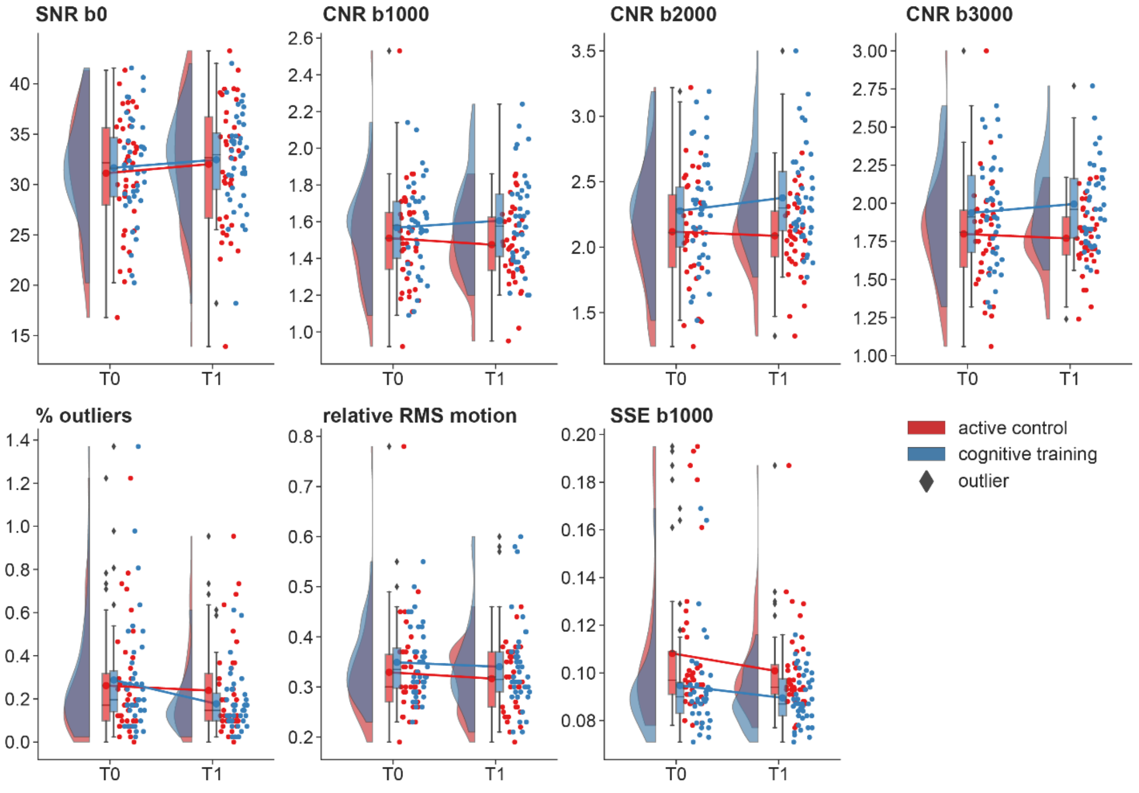


**Supplementary Figure 1 – Image quality measures.** Raincloud plots of the image quality measures (IQMs) for both the cognitive training and active control condition at both time points. There was a significant group effect of CNR of the b2000 and b3000 shell and SSE of the b1000 shell (see text). Abbreviations: SNR = signal-to-noise ratio, CNR = Contrast-to-noise ratio, RMS = root mean square, SSE = sum of squared error.

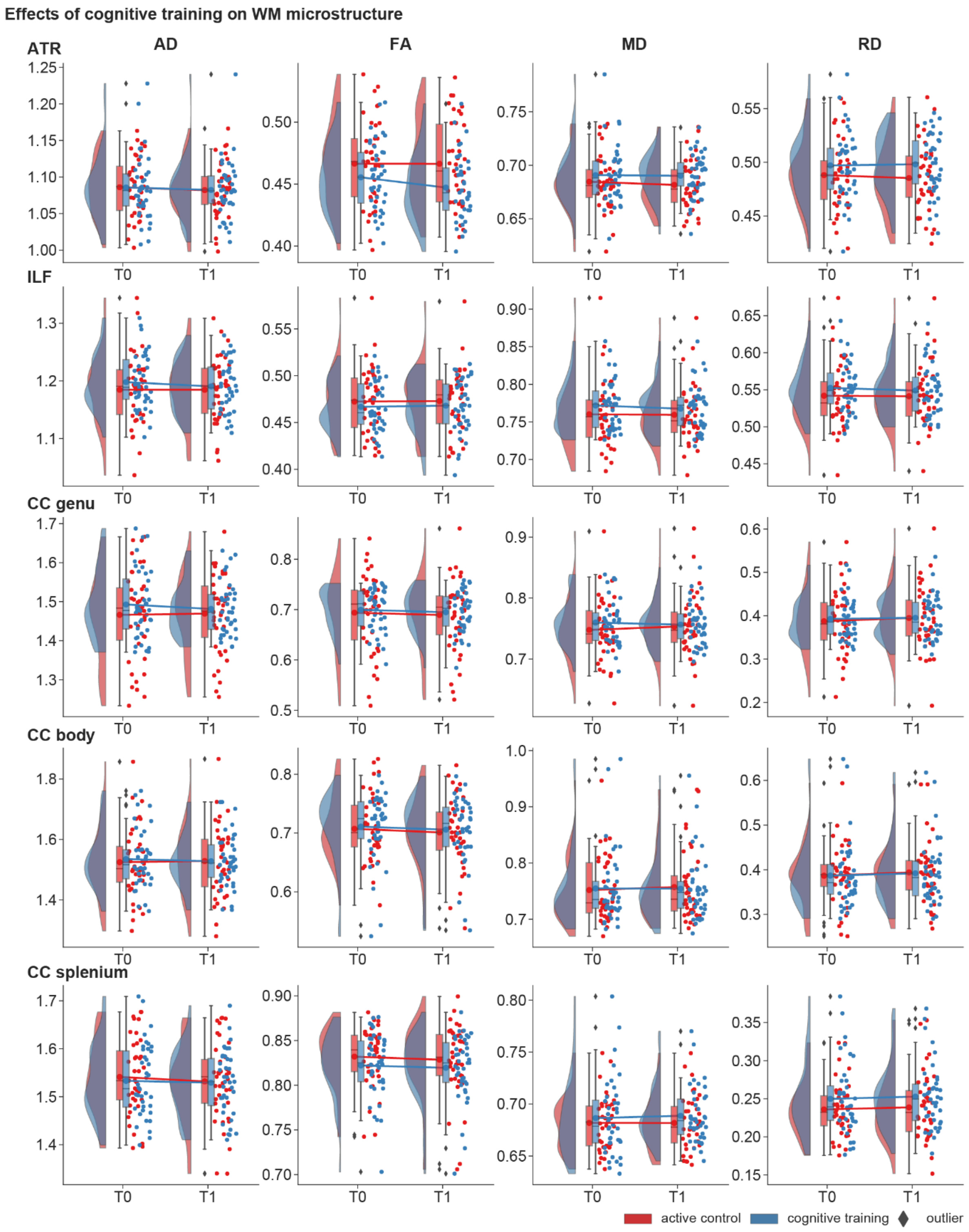


**Supplementary Figure 2 – Effects of cognitive training on WM microstructure.** Plots of the changes in axial diffusivity (AD), fractional anisotropy (FA), mean diffusivity (MD) and radial diffusivity (RD) in five different regions of interest the active control (red) and cognitive training (blue) group from pre-to-post intervention. Abbreviations: ATR = anterior thalamic radiation, ILF = inferior longitudinal fasciculus, CC = corpus callosum.

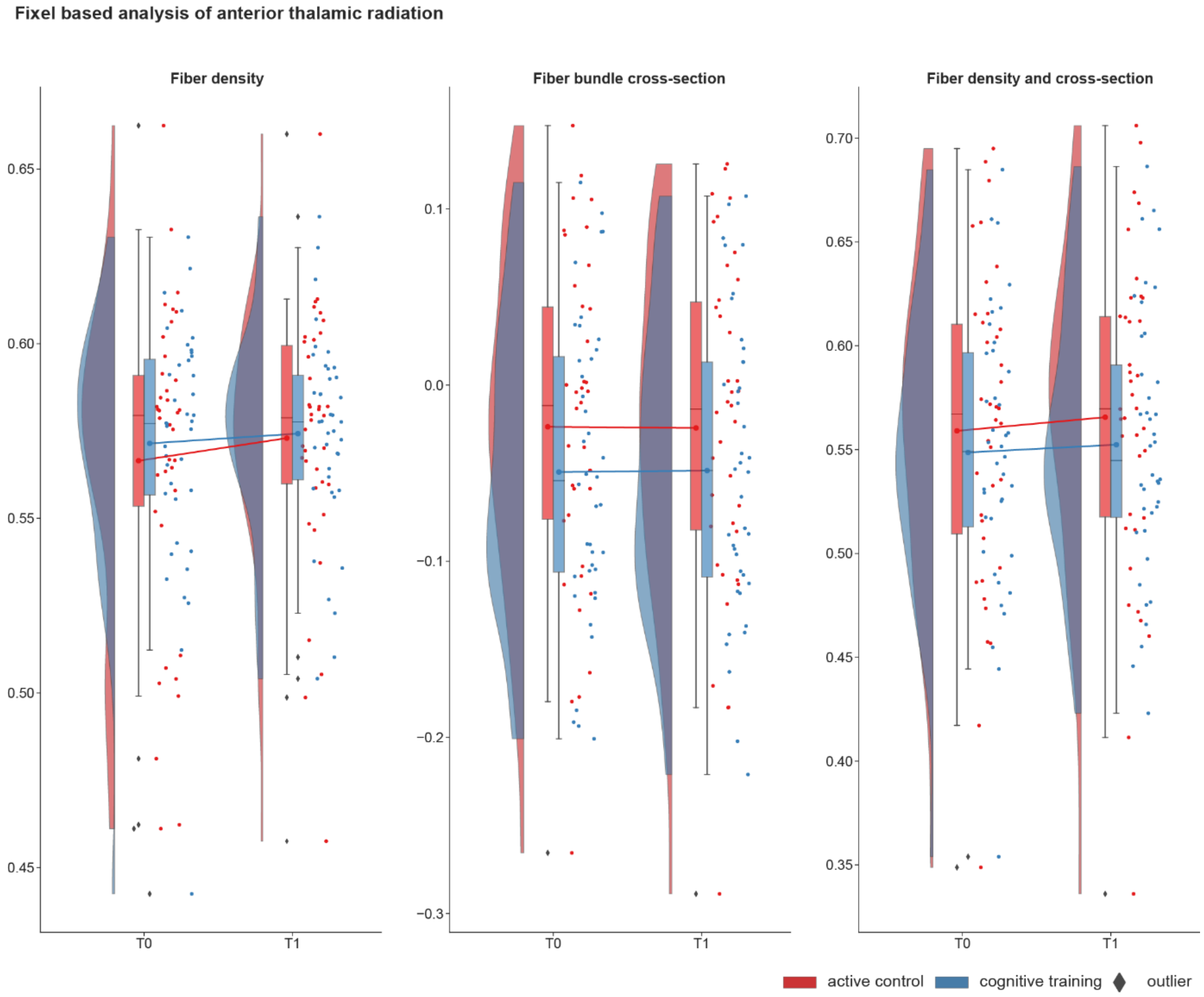


**Supplementary Figure 3 – effects of training on fixel based morphometry of the anterior thalamic radiation. .** Plots of the changes in the fiber density, fiber bundle cross-section and fiber bundle density and cross-section in the active control (red) and cognitive training (blue) group from pre-to-post intervention.

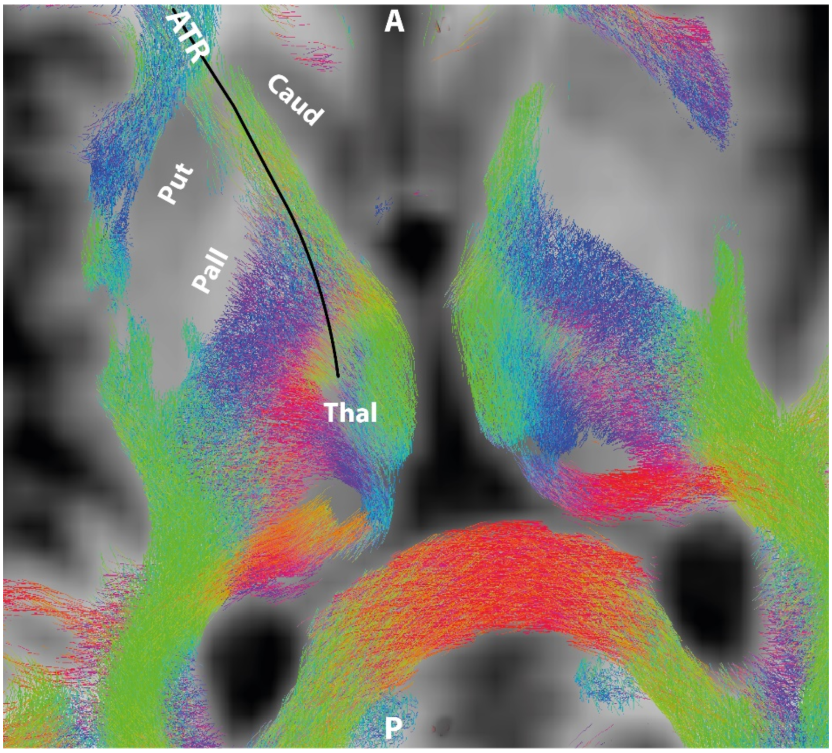


**Supplementary Figure 4 – tractogram overlaid with schematic trajectory of anterior thalamic radiation.** This figure shows that the trajectory of anterior thalamic radiation (ATR) from the prefrontal cortex to the thalamus (Thal), passing through the anterior limb of the internal capsule past the putamen (Put), pallidum (Pall), Caudate nucleus (Caud). Note the different colored fibers along the path of the ATR that signify different directions and the presence of crossing fibers in a large proportion of the voxels that make up the ATR.
