## Supplementary material for "Cognitive training in Parkinson’s disease induces local, not global, changes in white matter microstructure": COBIDAS checklist

Template generously supplied by Cassandra Gould van Praag, Department of Psychiatry, University of Oxford


| Reference | Table | Aspect1 | Aspect2 | Aspect3 | Aspect4 | Jan-19 | Mandatory | Data Source | Answer |
|---|---|---|---|---|---|---|---|---|---|
| D01.01.05.00.00.01 | Table D.1. Experimental Design Reporting | Number of subjects (by group) | Subjects participated and analyzed | | | CC.BY 4.0 | Y | Quality control log | see Figure 1 flowchart |
| D01.02.01.00.00.01 | Table D.1. Experimental Design Reporting | Inclusion criteria and descriptive statistics (by group) | Age | | | Mean, standard deviation and range | Y | Demographics | see Table 1 |
| D01.02.02.00.00.01 | Table D.1. Experimental Design Reporting | Inclusion criteria and descriptive statistics (by group) | Sex | | | Absolute counts or relative frequencies | Y | Demographics | see Table 1 |
| D01.02.04.00.00.01 | Table D.1. Experimental Design Reporting | Inclusion criteria and descriptive statistics (by group) | Education, SES | | | Education is essential for studies comparing patient and control groups; complete SES reporting less important for single- group studies, but still useful. Specify measurement instrument used; may be parental SES and education if study has minors | Y | Demographics | see Table 1 |
| D01.02.05.00.00.01 | Table D.1. Experimental Design Reporting | Inclusion criteria and descriptive statistics (by group) | IQ | | | Specify measurement instrument used | N | Demographics | not assessed |
| D01.02.06.00.00.01 | Table D.1. Experimental Design Reporting | Inclusion criteria and descriptive statistics (by group) | Handedness | | | Absolute or relative frequencies; basis of handedness-attribution (self report, EHI, other tests). (Important for fMRI, may be less important for structural studies) | Y | Demographics | not assessed |
| D01.02.07.00.00.01 | Table D.1. Experimental Design Reporting | Inclusion criteria and descriptive statistics (by group) | Exclusion criteria | | | Describe any screening criteria, including those applied to "normal" sample such as MRI exclusion criteria | Y | Protocol / Ethics | 1) a Montreal Cognitive Assessment score < 22, 2) indications of current drug- or alcohol abuse, 3) moderate to severe depressive symptoms, 4) an impulse control disorder, 5) psychotic symptoms except for benign hallucinations, or 6) a history of traumatic brain injury. Details are provided in (van Balkom et al. 2019). Exclusion criteria for participation in the MRI study were 1) a space occupying lesion, 2) significant vascular abnormalities (Fazekas > 1), 3) presence of metal in the body (e.g., a neurostimulator), 4) pregnancy, or 5) difficulty with, or shortness of breath during 60 minutes of lying still. |
| D01.02.08.00.00.01 | Table D.1. Experimental Design Reporting | Inclusion criteria and descriptive statistics (by group) | Clinical criteria | | | Detail the area of recruitment (in- vs. outpatient setting, community hospital vs. tertiary referral center etc.) as well as whether patients were currently in treatment | Y | Protocol / Ethics | diagnosed with Parkinson's disease and living in the Netherlands |
| D01.02.09.00.00.01 | Table D.1. Experimental Design Reporting | Inclusion criteria and descriptive statistics (by group) | Clinical instruments | | | Describe the instruments used to obtain the diagnosis and provide tests of intra- or inter--rater reliability. Clarify whether a "clinical diagnosis" or "inventory diagnosis" was used (if applicable). State the diagnostic system (ICD, DSM etc.) that was used | Y | Protocol | clinical diagnosis of Parkinson's disease according to UK Brain bank criteria. |
| D01.02.10.00.00.01 | Table D.1. Experimental Design Reporting | Inclusion criteria and descriptive statistics (by group) | Matching strategy | | | Describe how groups were matched | Y | Protocol | on age, sex and education level. See methods paper: https://doi.org/10.1186/s12883-019-1403-6 |
| D01.02.11.00.00.01 | Table D.1. Experimental Design Reporting | Inclusion criteria and descriptive statistics (by group) | Population & recruitment strategy | | | Population from which subjects were drawn, and how and where recruitment took place, e.g., schools, clinics, etc. If possible, note if subjects are research- naive or have participated in other studies before | Y | Protocol / Ethics | Parkinson's disease patients<br>General trial inclusion criteria were 1) mild to moderately advanced idiopathic PD (Hoehn & Yahr stage < 4), 2) significant subjective cognitive complaints (PD Cognitive Functional Rating Scale score > 3), access to and proficiency in using a computer or tablet with internet. General exclusion criteria were 1) a Montreal Cognitive Assessment score < 22, 2) indications of current drug- or alcohol abuse, 3) moderate to severe depressive symptoms (Beck Depression Inventory score > 18), 4) an impulse control disorder, 5) psychotic symptoms except for benign hallucinations, or 6) a history of traumatic brain injury. Exclusion criteria for participation in the MRI study were 1) presence of metal in the body (e.g., a neurostimulator), 2) pregnancy, 4) difficulty lying still for 60 minutes (e.g. due to shortness of breath), 5) a space occupying lesion or 6) significant vascular abnormalities (Fazekas > 1). more details in https://doi.org/10.1186/s12883-019-1403-6 |
| D01.02.12.00.00.01 | Table D.1. Experimental Design Reporting | Inclusion criteria and descriptive statistics (by group) | Subject scanning order | | | With multiple groups, information on ordering and or balance over time; especially report relative to scanner changes/upgrades. (Ideally, use randomized or interleaved order to avoid bias due to scanner changes/upgrades) | Y | Protocol | N/A |
| D01.02.13.00.00.01 | Table D.1. Experimental Design Reporting | Inclusion criteria and descriptive statistics (by group) | Neurocognitive measures | | | All measures collected on subjects should be described and reported | Y | Protocol | see https://doi.org/10.1186/s12883-019-1403-6 |

| | | | | | | | | | |
|---|---|---|---|---|---|---|---|---|---|
| D01.03.01.00.00.01 | Table D.1. Experimental Design Reporting | Ethical considerations | Ethical approval | | | Describe approval given, including the particular institutional review board, medical ethics committee or equivalent that granted the approval. When data is shared, describe the ethics/institutional approvals required from either the author (source) or recipient | Y | Ethics | medical ethical committee of VU University medical center (NL58750.029.16 |
| D01.03.02.00.00.01 | Table D.1. Experimental Design Reporting | Ethical considerations | Informed consent | | | Record whether subjects provided informed consent or, if applicable, informed assent | Y | Ethics | all participants provided written informed consent |
| D01.04.01.00.00.01 | Table D.1. Experimental Design Reporting | Design specifications | Design type | | | Task or resting state. Event- related or block design. (See body text for usage of 'block design' terminology) | Y | Protocol | N/A |
| D01.04.02.00.00.01 | Table D.1. Experimental Design Reporting | Design specifications | Condition & stimuli | | | Clearly describe each condition and the stimuli used. Be sure to completely describe baseline (e.g. blank white/black screen, presence of fixation cross, or any other text), especially for resting -state studies. When possible provide images or screen snapshots of the stimuli | Y | Protocol / Scanning task stimulus | N/A |
| D01.04.03.00.00.01 | Table D.1. Experimental Design Reporting | Design specifications | Number of blocks, trials or experimental units | | | Specify per session, and if differing by subject, summary statistics (mean, range and/or standard deviation) of such counts | Y | Protocol / Scanning task stimulus | N/A |
| D01.04.04.00.00.01 | Table D.1. Experimental Design Reporting | Design specifications | Timing and duration | | | Length of each trial or block (both, if trials are blocked), and interval between trials. Provide the timing structure of the events in the task, whether a random/jittered pattern or a regular arrangement; any jittering of block onsets | Y | Protocol / Scanning task stimulus | N/A |
| D01.04.05.00.00.01 | Table D.1. Experimental Design Reporting | Design specifications | Length of the experiment | | | Describe the total length of the scanning session, as well as the duration of each run. (Important to assess subject fatigue) | Y | Protocol | approximately 45 minutes for the entire scan session |
| D01.04.06.00.00.01 | Table D.1. Experimental Design Reporting | Design specifications | Design optimization | | | Whether design was optimized for efficiency, and how | Y | Protocol | N/A |
| D01.04.07.00.00.01 | Table D.1. Experimental Design Reporting | Design specifications | Presentation software | | | Name software, version and operating system on which the stimulus presentation was run. When possible, provide code used to drive experiment | Y | Scanner control room / Scanning task stimulus | N/A |
| D01.06.01.00.00.01 | Table D.1. Experimental Design Reporting | Power analysis | Outcome | | | The type of outcome used as the basis of power computations, e.g. signal in a pre-specified ROI, or whole image voxelwise (or cluster-wise, peak-wise, etc.) | Y | Protocol | N/A |
| D01.06.02.00.00.01 | Table D.1. Experimental Design Reporting | Power analysis | Power parameters | | | Effect size (or effect magnitude and standard deviation separately) | Y | Protocol | N/A |
| D01.06.02.00.00.02 | Table D.1. Experimental Design Reporting | Power analysis | Power parameters | | | Source of predicted effect size (previous literature with citation; pilot data with description, etc.) | Y | Protocol | N/A |
| D01.06.02.00.00.03 | Table D.1. Experimental Design Reporting | Power analysis | Power parameters | | | Significance level (e.g. uncorrected alpha 0.05 for an ROI, or FWE-corrected significance | Y | Protocol | N/A |
| D01.06.02.00.00.04 | Table D.1. Experimental Design Reporting | Power analysis | Power parameters | | | Target power (typically 80%) | Y | Protocol | N/A |
| D01.06.02.00.00.05 | Table D.1. Experimental Design Reporting | Power analysis | Power parameters | | | Any other parameters set (e.g. for spatial methods a brain volume and smoothness may be needed to be specified) | Y | Protocol | N/A |
| D01.07.01.00.00.01 | Table D.1. Experimental Design Reporting | Behavioral performance | Variables recorded | | | State number of type of variables recorded (e.g. correct button press, response time) | Y | Protocol | N/A |
| D01.07.02.00.00.01 | Table D.1. Experimental Design Reporting | Behavioral performance | Summary statistics | | | Summaries of behavior sufficient to establish that subjects were performing the task as expected. For example, correct response rates and/or response times, summarized over subjects (e.g. mean, range and/or standard deviation) | Y | Results | N/A |
| D02.01.01.00.00.01 | Table D.2. Acquisition Reporting | Subject preparation | Mock scanning | | | Use of an MRI simulator to acclimate subjects to scanner environment. Report type of mock scanner and protocol (i.e. duration, types of simulated scans, experiments) | N | Protocol | N/A |
| D02.01.02.00.00.01 | Table D.2. Acquisition Reporting | Subject preparation | Special accommodations | | | For example, for pediatric scanning, presence of parent/guardian in the room | Y | Protocol | N/A |
| D02.02.01.00.00.01 | Table D.2. Acquisition Reporting | MRI system description | Scanner | | | Provide make, model & field strength in tesla (T) | Y | Scanner control room | GE 3.0T Discovery MR750 (General Electronics, Milwaukee, US) |
| D02.02.02.00.00.01 | Table D.2. Acquisition Reporting | MRI system description | Coil | | | Receive coil (e.g. "a 12-channel phased array coil", but more details for a custom coil) and (if nonstandard) transmit coil. Additional information on the gradient system, e.g. gradient strength (if non-standard for the make and model, or switchable) | Y | Scanner control room | 32-channel head coil |
| D02.03.01.00.00.01 | Table D.2. Acquisition Reporting | MRI acquisition | Pulse sequence type | | | For example, gradient echo, spin echo, etc. | Y | Sequence protocol | single spin echo |
| D02.03.02.00.00.01 | Table D.2. Acquisition Reporting | MRI acquisition | Imaging type | | | For example, echo planar imaging (EPI), spiral, 3D | Y | Sequence protocol | EPI |
| D02.03.02.00.00.02 | Table D.2. Acquisition Reporting | MRI acquisition | Imaging type | | | Number of shots (if multi-shot); partial Fourier scheme & reconstruction method (if used) | Y | Sequence protocol | N/A |
| D02.03.03.01.00.01 | Table D.2. Acquisition Reporting | MRI acquisition | Essential sequence & imaging parameters | All acquisitions | | Echo time (TE) | Y | Sequence protocol | 81 ms |
| D02.03.03.01.00.02 | Table D.2. Acquisition Reporting | MRI acquisition | Essential sequence & imaging parameters | All acquisitions | | Repetition time (TR) | Y | Sequence protocol | 7350 ms |
| D02.03.03.01.00.03 | Table D.2. Acquisition Reporting | MRI acquisition | Essential sequence & imaging parameters | All acquisitions | | Flip angle (FA) | Y | Sequence protocol | 90 degrees |
| D02.03.03.01.00.04 | Table D.2. Acquisition Reporting | MRI acquisition | Essential sequence & imaging parameters | All acquisitions | | Acquisition time (duration of acquisition) | Y | Sequence protocol | 8 min 10 sec |
| D02.03.03.01.01.01 | Table D.2. Acquisition Reporting | MRI acquisition | Essential sequence & imaging parameters | All acquisitions | Multi-shot acquisitions | Time per volume | Y | Sequence protocol | - |

| | | | | | | | | | |
|---|---|---|---|---|---|---|---|---|---|
| D02.03.03.06.00.01 | Table D.2. Acquisition Reporting | MRI acquisition | Essential sequence & imaging parameters | Imaging parameters | | Field of view | Y | Sequence protocol | 240 mm |
| D02.03.03.06.00.02 | Table D.2. Acquisition Reporting | MRI acquisition | Essential sequence & imaging parameters | Imaging parameters | | In-plane matrix size, slice thickness and interslice gap, for 2D acquisitions | Y | Sequence protocol | 128x128, 2.5 mm slice thickness, no gap |
| D02.03.03.06.00.03 | Table D.2. Acquisition Reporting | MRI acquisition | Essential sequence & imaging parameters | Imaging parameters | | 3D matrix size, for 3D acquisitions | Y | Sequence protocol | - |
| D02.03.03.06.01.01 | Table D.2. Acquisition Reporting | MRI acquisition | Essential sequence & imaging parameters | Imaging parameters | Slice orientation | Axial, sagittal, coronal or oblique | Y | Sequence protocol | Axial |
| D02.03.03.06.01.02 | Table D.2. Acquisition Reporting | MRI acquisition | Essential sequence & imaging parameters | Imaging parameters | Slice orientation | Angulation: If acquisition not aligned with scanner axes, specify angulation to AC-PC line (see Slice position procedure) | Y | Sequence protocol | HYFA = hypophysis - fastigium line |
| D02.03.04.00.00.01 | Table D.2. Acquisition Reporting | MRI acquisition | Phase encoding | | | Specify phase encoding direction (e.g. as A/P, L/R, or S/I) | Y | Sequence protocol | A/P |
| D02.03.04.00.00.02 | Table D.2. Acquisition Reporting | MRI acquisition | Phase encoding | | | For 3D, specify “partition encode” (aka slice) direction | Y | Sequence protocol | - |
| D02.03.04.00.00.03 | Table D.2. Acquisition Reporting | MRI acquisition | Phase encoding | | | Phase encoding reversal: Mention if used (aka “blip-up/blip-down”) | Y | Sequence protocol | yes, blip-up/blip-down scans were acquired |
| D02.03.05.00.00.01 | Table D.2. Acquisition Reporting | MRI acquisition | Parallel imaging method & parameters | | | Method, e.g. SENSE, GRAPPA or other parallel imaging method, and acceleration factor | Y | Sequence protocol | parallel imaging factor = 2 |
| D02.03.05.00.00.02 | Table D.2. Acquisition Reporting | MRI acquisition | Parallel imaging method & parameters | | | Matrix coil mode, and coil combining method (if non-standard) | Y | Sequence protocol | - |
| D02.03.06.00.00.01 | Table D.2. Acquisition Reporting | MRI acquisition | Multiband parameters | | | Multiband factor and field-of-view shift (only if applicable) | Y | Sequence protocol | N/A |
| D02.03.07.00.00.01 | Table D.2. Acquisition Reporting | MRI acquisition | Readout parameters | | | Receiver bandwidth, readout duration, echo spacing | N | Sequence protocol | total readouttime= 0.028120 |
| D02.03.08.00.00.01 | Table D.2. Acquisition Reporting | MRI acquisition | Fat suppression | | | For anatomical scans, whether it was used or not | Y | Sequence protocol | for T1w MPRAGE scan yes |
| D02.03.09.00.00.01 | Table D.2. Acquisition Reporting | MRI acquisition | Shimming | | | Any specialized shimming procedures | Y | Sequence protocol | HOS |
| D02.03.12.00.00.01 | Table D.2. Acquisition Reporting | MRI acquisition | Brain coverage | | | Report whether coverage was whole-brain, and whether cerebellum and brainstem were included. If not whole-brain, note the nature of the partial area of coverage. If axial and co-planar with AC-PC line, the volume coverage in terms of Z in mm | Y | Sequence protocol | whole-brain but not every subject had full coverage of the cerebellum |
| D02.03.13.00.00.01 | Table D.2. Acquisition Reporting | MRI acquisition | Scanner-side preprocessing | | | Reconstruction matrix size differing from acquisition matrix size | Y | Sequence protocol | No |
| D02.03.13.00.00.02 | Table D.2. Acquisition Reporting | MRI acquisition | Scanner-side preprocessing | | | Prospective-motion correction (including details of any optical tracking, and how motion parameters are used) | Y | Sequence protocol | N/A |
| D02.03.13.00.00.03 | Table D.2. Acquisition Reporting | MRI acquisition | Scanner-side preprocessing | | | Signal inhomogeneity correction | Y | Sequence protocol | intensity non-uniformity correction for 3D T1-weighted structural image (MPRAGE) |
| D02.03.13.00.00.04 | Table D.2. Acquisition Reporting | MRI acquisition | Scanner-side preprocessing | | | Distortion-correction | Y | Sequence protocol | 3D geometric distortion correction for 3D T1-weighted structural image (MPRAGE) |
| D02.03.14.00.00.01 | Table D.2. Acquisition Reporting | MRI acquisition | Scan duration | | | In seconds | N | Sequence protocol | |
| D02.03.16.00.00.01 | Table D.2. Acquisition Reporting | MRI acquisition | T1 stabilization | | | Number of initial “dummy” scans acquired and then discarded by the scanner | Y | Sequence protocol | N/A |
| D02.04.01.00.00.01 | Table D.2. Acquisition Reporting | Preliminary quality control | Motion monitoring | | | For functional or diffusion acquisitions, any visual or quantitative checks for severe motion; likewise, for structural images, checks on motion or general image quality | Y | Protocol | N/A |
| D02.04.02.00.00.01 | Table D.2. Acquisition Reporting | Preliminary quality control | Incidental findings | | | Protocol for review of any incidental findings, and how they are handled in particular with respect to possible exclusion of a subject’s data | N | Protocol | all scans were reviewed by neuroradiologist. None had to be excluded |
| D03.01.00.00.00.01 | Table D.3. Preprocessing Reporting | Software | | | | For each software used, be sure to include version and revision number | Y | Analysis plan | fmriprep v1.4.0, fsl 6.0.1., MRtrix3.0.0, DTI-TK 2.3.1 |
| D03.02.00.00.00.01 | Table D.3. Preprocessing Reporting | Software citation | | | | Include URL and Research Resource Identifier for each software used | N | Analysis plan | fmriprep.org<br>https://fsl.fmrib.ox.ac.uk/fsl/fslwiki<br>https://www.mrtrix.org/<br>http://dti-tk.sourceforge.net/pmwiki/pmwiki.php<br>https://www.github.com/chrisvriend/DWI_processing_COGTIPS |
| D03.03.00.00.00.01 | Table D.3. Preprocessing Reporting | T1 stabilization | | | | Number of initial “dummy” scans discarded as part of preprocessing (if not already performed by scanner) | Y | Analysis plan | N/A |
| D03.04.00.00.00.01 | Table D.3. Preprocessing Reporting | Brain extraction | | | | Name of software/method (e.g., BET, recon-all in FreeSurfer, etc.) | Y | Analysis plan | fmriprep v1.4.0 (= ANTS + FreeSurfer 6.0.1) |
| D03.04.00.00.00.02 | Table D.3. Preprocessing Reporting | Brain extraction | | | | Parameter choices (e.g. BET’s fractional intensity threshold) | Y | Analysis plan | standard fmriprep v1.4.0 settings |
| D03.04.00.00.00.03 | Table D.3. Preprocessing Reporting | Brain extraction | | | | Any manual editing applied to the brain masks | Y | Analysis log | N/A |
| D03.05.00.00.00.01 | Table D.3. Preprocessing Reporting | Segmentation | | | | For structural images, method used to extract gray, white, CSF and other tissue classes | Y | Analysis plan | FSL FAST within fmriprep v1.4.0 |
| D03.07.00.00.00.01 | Table D.3. Preprocessing Reporting | Motion correction | | | | Name of software/method | Y | Analysis plan | with eddy in FSL 6.0.1 |
| D03.07.00.00.00.02 | Table D.3. Preprocessing Reporting | Motion correction | | | | Use of non-ridged registration, and if so the type of transformation | Y | Analysis plan | yes, FSL epi_reg BBR for T1 - DWI |
| D03.07.00.00.00.03 | Table D.3. Preprocessing Reporting | Motion correction | | | | Use of motion susceptibility correction (fieldmap -based unwarping), as well as the particular software/method | Y | Analysis plan | yes, blip-up/blip-down scans were acquired and used to correct for susceptibility correction using topup and eddy in FSL 6.0.1 |
| D03.07.00.00.00.04 | Table D.3. Preprocessing Reporting | Motion correction | | | | Reference scan (e.g. 1st scan or middle scan) | Y | Analysis plan | 1st scan; see Andersson et al. 2016 Neuroimage 125: 1063-78. |
| D03.07.00.00.00.05 | Table D.3. Preprocessing Reporting | Motion correction | | | | Image similarity metric (e.g. normalized correlation, mutual information, etc.) | Y | Analysis plan | N/A; see Andersson et al. 2016 Neuroimage 125: 1063-78. |

| | | | | | | | |
|---|---|---|---|---|---|---|---|
| D03.07.00.00.00.06 | Table D.3. Preprocessing Reporting | Motion correction | | Interpolation type (e.g., spline, sinc), and whether image transformations are combined to allow a single interpolation | Y | Analysis plan | cubic splines; see Andersson et al. 2016 Neuroimage 125: 1063-78. |
| D03.07.00.00.00.07 | Table D.3. Preprocessing Reporting | Motion correction | | Use of any slice--to--volume registration methods, or integrated with slice time correction | Y | Analysis plan | slice-to-volume registration: Andersson et al 2017, Neuroimage, 152, 450-66 |
| D03.08.00.00.00.01 | Table D.3. Preprocessing Reporting | Gradient distortion correction | | (If not already described as part of motion susceptibility correction) | Y | Analysis plan | |
| D03.09.00.00.00.01 | Table D.3. Preprocessing Reporting | Diffusion MRI eddy current correction | | Name of software/method, and if integrated with motion correction | Y | Analysis plan | FSL eddy implemented in fsl 6.0.1 |
| D03.09.00.00.00.02 | Table D.3. Preprocessing Reporting | Diffusion MRI eddy current correction | | Image similarity / cost function | Y | Analysis plan | FSL eddy default |
| D03.09.00.00.00.03 | Table D.3. Preprocessing Reporting | Diffusion MRI eddy current correction | | Type of transformation (e.g. rigid body, affine) and whether constrained only along the phase encode direction | Y | Analysis plan | FSL eddy default |
| D03.09.00.00.00.04 | Table D.3. Preprocessing Reporting | Diffusion MRI eddy current correction | | Note if gradient table (b-matrix) is then re-oriented | Y | Analysis plan | yes |
| D03.09.00.00.00.05 | Table D.3. Preprocessing Reporting | Diffusion MRI eddy current correction | | Volumetric change applied for eddy current along the phase--encode axis (by the Jacobian determinant) | Y | Analysis plan | no |
| D03.10.00.00.00.01 | Table D.3. Preprocessing Reporting | Diffusion estimation | | For all methods, report Model, parameterization and number of free parameters | Y | Analysis plan | FSL DTI-FIT default |
| D03.10.00.00.00.02 | Table D.3. Preprocessing Reporting | Diffusion estimation | | For all methods, report Estimation method | Y | Analysis plan | FSL DTI-FIT default |
| D03.10.00.00.00.03 | Table D.3. Preprocessing Reporting | Diffusion estimation | | For all methods, report Outlier handling approach | Y | Analysis plan | Outlier replacement implemented in FSL eddy (in fsl 6.0.1) |
| D03.10.00.00.00.04 | Table D.3. Preprocessing Reporting | Diffusion estimation | | For all methods, report Some evidence of fit quality; e.g. sample of slices of diffusion weighted data, or residual maps | Y | Analysis plan | eddyqc was performed and image quality measures, including sum of squared errors are reported in the manuscript |
| D03.10.01.00.00.01 | Table D.3. Preprocessing Reporting | Diffusion estimation | Tensor or Kurtosis | For Tensor or Kurtosis: Any parameter constraints, like cylindrical symmetry | Y | Analysis plan | Tensor |
| D03.10.02.00.00.01 | Table D.3. Preprocessing Reporting | Diffusion estimation | Multi-compartmental models | Compartments of the model | Y | Analysis plan | GM, WM, CSF - multishell multitissue |
| D03.10.03.00.00.01 | Table D.3. Preprocessing Reporting | Diffusion estimation | Orientation distribution function | Parametric (model) or nonparametric (basis function) model | Y | Analysis plan | msmt_5tt in MRtrix3 for tractography analysis |
| D03.10.03.00.00.02 | Table D.3. Preprocessing Reporting | Diffusion estimation | Orientation distribution function | Whether orientation distribution function or fibre orientation density is reported | Y | Analysis plan | ODF |
| D03.10.03.00.00.03 | Table D.3. Preprocessing Reporting | Diffusion estimation | Orientation distribution function | For spherical deconvolution, note how the canonical fibre response function is derived (e.g. from the data themselves, or simulated data) | Y | Analysis plan | for tractography: Multi-shell multi-tissue contrained spherical deconvolution on subject data |
| D03.11.00.00.00.01 | Table D.3. Preprocessing Reporting | Diffusion processing | | Summary measures computed (FA, MD, AD, RD, MK, AK, RK, etc.) | N | Analysis plan | for microstructure analyses: FA, MD, AD, RD for fixel-based analyses: fiber density, fiber cross-section and fiber density & cross-section |
| D03.11.00.00.00.02 | Table D.3. Preprocessing Reporting | Diffusion processing | | Whether a track based or voxel--wise method is used | N | Analysis plan | track-based for microstructure analyses with DTI-TK |
| D03.11.00.00.00.03 | Table D.3. Preprocessing Reporting | Diffusion processing | | Threshold used to define analysis voxels | N | Analysis plan | |
| D03.11.00.00.00.04 | Table D.3. Preprocessing Reporting | Diffusion processing | | Use of population reference track atlas vs. custom atlas (specify set of subjects used to create atlas) | N | Analysis plan | created group atlas for TBSS; see DTI-TK pipeline on https://www.github.com/chrisvriend/DWI_processing_COGTIPS |
| D03.11.00.00.00.05 | Table D.3. Preprocessing Reporting | Diffusion processing | | Standard deviation map (across subjects) | N | Analysis plan | no |
| D03.12.00.00.00.01 | Table D.3. Preprocessing Reporting | Diffusion tractography | | Name of software/method | Y | Analysis plan | MRtrix3 |
| D03.12.00.00.00.02 | Table D.3. Preprocessing Reporting | Diffusion tractography | | Step size, turning angle and stopping criteria | Y | Analysis plan | MRTRIX3 defaults: steplength = (iFOD2:) 0.5 x voxelsize, turning angle: 45 degrees. stopping criteria: tckgen will stop when the number of seeds (100M) attempted reaches the number specified here, |
| D03.12.00.00.00.03 | Table D.3. Preprocessing Reporting | Diffusion tractography | | For ROI based analysis, definition of ROIs (e.g. specify the images used to draw ROIs; manual, semi-automatic or automatic definition of ROIs) | Y | Analysis plan | N/A |
| D03.12.00.00.00.04 | Table D.3. Preprocessing Reporting | Diffusion tractography | | For tracking, note step--size, turning angle, any anatomical constraints imposed, and stopping criteria | Y | Analysis plan | see above |
| c | Table D.3. Preprocessing Reporting | Diffusion tractography | | If a measure of path probability / "connectivity" is extracted, clearly define this measure | Y | Analysis plan | connectivity = number of streamlines after correction with SIFT2 |
| D03.15.00.00.00.01 | Table D.3. Preprocessing Reporting | Distortion correction | | Use of any distortion correction due to field or gradient nonlinearity | Y | Analysis plan | FSL topup/eddy |
| D03.16.00.00.00.01 | Table D.3. Preprocessing Reporting | Intersubject registration | | Name of software/method (e.g., FSL flirt followed by fnirt, FreeSurfer, Caret, Workbench, etc.) | Y | Analysis plan | FSL epi_reg |
| D03.16.00.00.00.02 | Table D.3. Preprocessing Reporting | Intersubject registration | | Whether volume and/or surface based registration is used (if not already clearly implied) | Y | Analysis plan | volume-based |
| D03.16.00.00.00.03 | Table D.3. Preprocessing Reporting | Intersubject registration | | Image types registered (e.g. T2* or T1) | Y | Analysis plan | DWI scans are non-linearly registered using DTI-TK |
| D03.16.00.00.00.04 | Table D.3. Preprocessing Reporting | Intersubject registration | | Any preprocessing to images; e.g. for T1, bias field correction, or segmentation of gray matter; for T2*, single image (specify image) or mean image | Y | Analysis plan | dependent on modality: see scripts on https://www.github.com/chrisvriend/DWI_processing_COGTIPS |

| | | | | | | | | |
|---|---|---|---|---|---|---|---|---|
| D03.16.00.00.00.05 | Table D.3. Preprocessing Reporting | Intersubject registration | | | Template space (e.g., MNI, Talairach, fsaverage, FS_LR), modality (e.g., T1, T2*), resolution (e.g., 2mm, fsaverage5, 32k_FS_LR), and the specific name of template image used; note the domain of the template if not whole brain, i.e. cortical surface only, cerebellum only, CIFTI 'grayordinates' (cortical surface vertices + subcortical gray matter voxels), etc. | Y | Analysis plan | atlas of WM tracts in MNI space and subsequently warped to the group template; see https://www.github.com/chrisvriend/DWI_processing_COGTIPS |
| D03.16.00.00.00.06 | Table D.3. Preprocessing Reporting | Intersubject registration | | | Additional template transformation for reporting; e.g., if using a template in MNI space, but reporting coordinates in Talairach, clearly note and report method used (e.g., Brett's mni2tal, Lancaster's icbm_spm2tal) | Y | Analysis plan | N/A |
| D03.16.00.00.00.07 | Table D.3. Preprocessing Reporting | Intersubject registration | | | Choice of warp (rigid, nonlinear); if nonlinear, transformation type (e.g., B--splines, stationary velocity field, momentum, non-parametric displacement field); if a parametric transformation is used, report resolution, e.g., 10x10x10 spline control points | Y | Analysis plan | DTI-TK: diffeomorphic<br>fixel analysis in MRtrix3: linear with subsequent non-linear registration to to create a group template (population_template) |
| D03.16.00.00.00.08 | Table D.3. Preprocessing Reporting | Intersubject registration | | | Use of regularization, and the parameter(s) used to set degree of regularization | Y | Analysis plan | DTI-TK and MRtrix3 default settings |
| D03.16.00.00.00.09 | Table D.3. Preprocessing Reporting | Intersubject registration | | | Interpolation type (e.g., spline, linear); if projection from volume to surface space, how were voxels sampled from the volume (e.g., trilinear; nearest neighbor; ribbon--constrained specifying inner and outer surface used). | Y | Analysis plan | DTI-TK and MRtrix3 default settings |
| D03.16.00.00.00.10 | Table D.3. Preprocessing Reporting | Intersubject registration | | | Cost function (e.g., correlation ratio, mutual information, SSD) | Y | Analysis plan | DTI-TK and MRtrix3 default settings |
| D03.16.00.00.00.11 | Table D.3. Preprocessing Reporting | Intersubject registration | | | Use of cost--function masking | Y | Analysis plan | DTI-TK and MRtrix3 default settings |
| D03.17.00.00.00.01 | Table D.3. Preprocessing Reporting | Intensity correction | | | Bias field corrections for structural MRI, but also correction of odd versus even slice intensity differences attributable to interleaved EPI acquisition without gaps | Y | Analysis plan | bias field correction of T1w as part of freesurfer 6.0.1 |
| D03.18.00.00.00.01 | Table D.3. Preprocessing Reporting | Intensity normalization | | | Scan-by-scan or run-wide scaling of image intensities before statistical modelling. E.g. SPM scales each run such that the mean image will have mean intracerebral intensity of 100; FSL scales each run such that the mean image will have an intracerebral mode of 10,000 | N | Analysis plan | N/A |
| D03.19.01.00.00.01 | Table D.3. Preprocessing Reporting | Artifact and structured noise removal | Physiological noise correction method | | Name of software/method used (e.g. CompCor, ICA-FIX, ICA-AROMA, etc.) | Y | Analysis plan | N/A |
| D03.19.02.01.00.01 | Table D.3. Preprocessing Reporting | Artifact and structured noise removal | Nuisance regression | Motion parameters | Expansion basis and order (e.g. 1st temporal derivatives; Volterra kernel expansion) | Y | Analysis plan | N/A |
| D03.19.02.02.00.01 | Table D.3. Preprocessing Reporting | Artifact and structured noise removal | Nuisance regression | Tissue signals | Tissue type (e.g., whole brain, gray matter, white matter, ventricles) | Y | Analysis plan | N/A |
| D03.19.02.02.00.02 | Table D.3. Preprocessing Reporting | Artifact and structured noise removal | Nuisance regression | Tissue signals | Tissue definition (e.g., a priori seed, automatic segmentation, spatial regression) | Y | Analysis plan | N/A |
| D03.19.02.02.00.03 | Table D.3. Preprocessing Reporting | Artifact and structured noise removal | Nuisance regression | Tissue signals | Signal definition (e.g., mean of voxels, first singular vector, etc.) | Y | Analysis plan | N/A |
| D03.19.02.03.00.01 | Table D.3. Preprocessing Reporting | Artifact and structured noise removal | Nuisance regression | Physiological signals | e.g., heart rate variability, respiration | Y | Analysis plan | N/A |
| D03.19.02.03.00.02 | Table D.3. Preprocessing Reporting | Artifact and structured noise removal | Nuisance regression | Physiological signals | Modeling choices (e.g. RETROICOR, cardiac and/or respiratory response functions) and number of computed regressors | Y | Analysis plan | N/A |
| D03.20.00.00.00.01 | Table D.3. Preprocessing Reporting | Volume censoring (a.k.a. "scrubbing" or "de-spiking") | | | Name of software/method | Y | Analysis plan | N/A |
| D03.20.00.00.00.02 | Table D.3. Preprocessing Reporting | Volume censoring (a.k.a. "scrubbing" or "de-spiking") | | | Criteria (e.g., frame-by-frame displacement threshold, percentage BOLD change) | Y | Analysis plan | N/A |
| D03.20.00.00.00.03 | Table D.3. Preprocessing Reporting | Volume censoring (a.k.a. "scrubbing" or "de-spiking") | | | Use of censoring or interpolation; if interpolation, method used (e.g., spline, spectral estimation) | Y | Analysis plan | N/A |
| D03.23.00.00.00.01 | Table D.3. Preprocessing Reporting | Quality control reports | | | Summaries of subject motion (e.g. mean frame-wise displacement), image variance (e.g. DVARS), and note of any other irregularities found (e.g. motion or poor SNR not sufficiently severe to warrant exclusion). Should be included with any publically shared data | N | Results | see Figure 2 and results section |
| D04.01.01.01.00.01 | Table D.4. Statistical Modeling & Inference | Mass univariate analyses | Dependent variable | Data submitted to statistical modeling | Report the number of time points, number of subjects; specify exclusions of time points / subjects, if not already specified in experimental design | Y | Analysis log | see Table 1 |
| D04.01.01.02.00.01 | Table D.4. Statistical Modeling & Inference | Mass univariate analyses | Dependent variable | Spatial region modeled | If not "Full brain", give a specification of an anatomically or functionally defined mask | Y | Analysis plan | whole-brain but not every subject had full coverage of the cerebellum |
| D04.03.01.00.00.01 | Table D.4. Statistical Modeling & Inference | Multivariate modelling & predictive analysis | Independent variables | | Variable type (discrete or continuous) | Y | Analysis plan | N/A |
| D04.03.01.00.00.02 | Table D.4. Statistical Modeling & Inference | Multivariate modelling & predictive analysis | Independent variables | | Class proportions in classification settings | Y | Analysis plan | N/A |
| D06.01.01.00.00.01 | Table D.6. Data Sharing | Reporting a data sharing resource | Material shared | | List types of images and non-imaging data provided | Y | Data management plan | N/A |
| D06.01.01.00.00.02 | Table D.6. Data Sharing | Reporting a data sharing resource | Material shared | | Report on the completeness of the data (e.g., number of subjects where all types of imaging, demographic, and behavioral data is available) | Y | Results | N/A |
| D06.01.02.00.00.01 | Table D.6. Data Sharing | Reporting a data sharing resource | URL, access information | | Stable URL or DOI | Y | Data management plan | N/A |

| | | | | | | | |
|---|---|---|---|---|---|---|---|
| D06.01.02.00.00.02 | Table D.6. Data Sharing | Reporting a data sharing resource | URL, access information | Specific instructions on how to gain access. Specifically mention whether application must be vetted for particular intended research use (e.g. to preclude multiple users investigating the same question), or whether a research collaboration must be established | Y | Data management plan | N/A |
| D06.01.02.00.00.03 | Table D.6. Data Sharing | Reporting a data sharing resource | URL, access information | Cost of access | Y | Data management plan | N/A |
| D06.01.03.00.00.01 | Table D.6. Data Sharing | Reporting a data sharing resource | Ethics compliance | Confirm that the ethics board of the host institution generating the data approves the sharing of the data made available | Y | Ethics | yes but with certain restrictions |
| D06.01.03.00.00.02 | Table D.6. Data Sharing | Reporting a data sharing resource | Ethics compliance | Clarify any constraints on uses of shared data, for example, whether users downloading the data also need ethics approval from their own institution | Y | Ethics | data can be requested from the authors upon reasonable request, data sharing agreement first needs to be set up by legal departments of both institutes |
| D07.01.13.00.00.01 | Table D.7. Reproducibility | Documentation | Tools used | Tool names, versions, and URLs | Y | Analysis plan | fmriprep v1.4.0 - fmriprep.org<br>fsl 6.0.1. https://fsl.fmrib.ox.ac.uk/fsl/fslwiki<br>MRtrix3.0 https://www.mrtrix.org/<br>DTI-TK 2.3.1 http://dti-tk.sourceforge.net/pmwiki/pmwiki.php<br>pipeline used - https://www.github.com/chrisvriend/DWI_processing_COGTIPS |
| D07.02.01.00.00.01 | Table D.7. Reproducibility | Archiving | Tools availability | Note if tools are publically available | N | Analysis plan | yes |
| D07.02.02.00.00.01 | Table D.7. Reproducibility | Archiving | Virtual appliances | Note if a virtual environment to facilitate a repeated analysis is available | N | Data management plan | no |
| D07.03.01.00.00.01 | Table D.7. Reproducibility | Data | Data | Provide permanent identifier if possible | N | Data management plan | N/A |
| D07.04.01.00.00.01 | Table D.7. Reproducibility | Workflow | Workflow | Provide permanent identifier if possible | N | Data management plan | N/A |
